# Scaling Self-Supervised Learning for Histopathology with Masked Image Modeling

**DOI:** 10.1101/2023.07.21.23292757

**Authors:** Alexandre Filiot, Ridouane Ghermi, Antoine Olivier, Paul Jacob, Lucas Fidon, Axel Camara, Alice Mac Kain, Charlie Saillard, Jean-Baptiste Schiratti

## Abstract

Computational pathology is revolutionizing the field of pathology by integrating advanced computer vision and machine learning technologies into diagnostic workflows. It offers unprecedented opportunities for improved efficiency in treatment decisions by allowing pathologists to achieve higher precision and objectivity in disease classification, tumor microenvironment description and identification of new biomarkers. However, the potential of computational pathology in personalized medicine comes with significant challenges, particularly in annotating whole slide images (WSI), which is time-consuming, costly and subject to inter-observer variability. To address these challenges, Self-Supervised Learning (SSL) has emerged as a promising solution to learn representations from histology patches and leverage large volumes of unlabelled WSI. Recently, Masked Image Modeling (MIM) as a SSL framework has emerged and is now considered to outperform purely contrastive learning paradigms. In this work, we therefore explore the application of MIM to histology using iBOT, a self-supervised transformer-based framework. Through a wide range of 17 downstream tasks over seven cancer indications, both at the slide and patch levels, we provide recommendations on the pre-training of large models for histology data using MIM. First, we demonstrate that in-domain pre-training with iBOT outperforms both ImageNet pre-training and a model pre-trained with a purely contrastive learning objective, MoCo v2. Second, we show that Vision Transformers (ViT) models, when scaled appropriately, have the capability to learn pan-cancer representations that benefit a large variety of downstream tasks. Finally, our iBOT ViT-Base model (80 million parameters), pre-trained on more than 40 million histology images from 16 different cancer types, achieves state-of-the-art performance in most weakly-supervised WSI classification tasks compared to other SSL frameworks available in the literature. This paves the way for the development of a foundation model for histopathology. Our code, models and features are publicly available at https://github.com/owkin/HistoSSLscaling.

## 1 Introduction

Histopathology plays a crucial role in disease diagnosis, treatment planning, and medical research. In clinical routine workflows, pathologists analyze histology slides manually to identify cellular abnormalities, tissue patterns and disease markers. Computational pathology has brought a paradigm shift in how histology is approached, leveraging advanced technologies such as Deep Learning to enhance accuracy, efficiency, and reproducibility in the analysis of histological images. The application of computational pathology is revolutionizing pathology, transforming the way diseases are detected, classified and treated (1). Additionally, computational pathology offers the potential to quantify tumor microenvironments, discover new biomarkers and improve patient and disease stratification (2, 3).

In the last decade, Deep Learning has made significant progress in medical image analysis. These advancements have enabled researchers to leverage massive amounts of annotated datasets to improve patient outcomes (4) and integrate artificial intelligence-based solutions into clinical workflows (5). However, labeling large amounts of data can be challenging, especially when dealing with medical data. Indeed, annotating WSI at the slide or pixel level can be tedious and time-consuming for trained pathologists. Moreover, the diversity of cancer types and tissue preparation protocols further complicates the annotation task as it likely introduces variability in color, texture, staining, and cellular morphology.

Motivated by the lack of large-scale annotated datasets, the field of computer-aided medical imaging has witnessed a widespread adoption of transfer learning from ImageNet (6). A number of studies have successfully applied transfer learning to digital pathology (7–11). As a matter of fact, convolutional neural networks (CNN) trained on the ImageNet database have learnt robust visual representations from natural images and serve as powerful feature extractors for histology images. However, relying solely on out-of-domain pre-training such as ImageNet has limitations, particularly due to domain shift, lower color variation and no canonical orientation (12). Histology images exhibit complex and specific features, including cellular structures, tissue morphology and staining patterns, which may not be adequately captured by models pre-trained on ImageNet (13).

In recent years, SSL methods have made spectacular progress on ImageNet, bridging the gap with fully-supervised methods and eliminating the need for labeled data. SSL methods allow learning relevant representations from unlabeled images by formulating and solving a pretext task (14). Recently, these methods have been used to leverage vast amounts of unlabeled WSI and perform unsupervised feature learning (13, 15–24). Recent studies have successfully applied and tailored existing SSL frameworks to histology images; see (25) for an extensive review. However, the majority of these studies have focused on small ViT (26) or CNN models pre-trained with self-distillation and contrastive learning (CL), especially through DINO (27) or MoCo v2 (28). In this work, we show that leveraging more recent advances in SSL, especially masked image modeling with ViT models, is beneficial for histopathology and outperforms both ImageNet pre-training and a model pre-trained with a purely contrastive learning objective, *e*.*g*., MoCo v2. Notably, we train a large ViT with more than 300M parameters, which is to the best of our knowledge the largest model ever trained on histology images.

Inspired by BERT (29) and Masked Language Modeling (MLM), MIM (26, 30) is another recent emerging SSL paradigm which has become popular due to its impressive fine-tuning performance on a variety of downstream computer vision tasks (30–33) and its robustness against image artifacts (33). Despite its potential in digital pathology, the application of MIM to histology data remains largely unexplored. Indeed, despite high fine-tuning capabilities, models pre-trained with MIM also exhibit poor linear probing performance (34), which is critical in digital pathology where most applications involve the aggregation of pre-extracted features for outcome prediction. By combining MIM and CL, the iBOT framework (image BERT pre-training with Online Tokenizer, (33)) addresses this limitation. iBOT takes advantage of the architecture of ViT, their effectiveness on computer vision tasks and performs self-distillation both on masked image patches to capture low-level details and on the class token to acquire high-level visual semantics. In addition to achieving state-of-the-art results in downstream tasks like classification or semantic segmentation, iBOT exhibits high fine-tuning and linear probing performance in low data regime, and robustness properties against various perturbations such as background change and occlusion. Those properties are highly relevant for digital pathology. Additionally, recent studies (35) have shown promising results regarding the scalability of MIM on ImageNet. Still, to the best of our knowledge, no previous study has investigated whether MIM pre-training on histology data can benefit from larger architectures and larger pre-training datasets, thereby establishing the possibility of a foundation model specifically for histology.

In this paper, we investigate the application of MIM to histology images using the ViT-based iBOT framework and provide insights on how to select the model architecture based on the amount of data available. Our main contributions can be summarized as follows:

- We assess the representation capability of iBOT through a large panel of 17 downstream tasks over seven cancer indications, covering both weakly-supervised WSI classification and supervised patch classification. These downstream tasks consist in predicting genomic alterations such as Microsatellite Instability (MSI) or Homologous Recombination Deficiency (HRD), histological and molecular subtypes classification, or overall survival (OS) prediction; See figure 1. All slide-level experiments were conducted through nested cross-validation;
- We demonstrate that in-domain pre-training with iBOT outperforms ImageNet pre-training on comparable model architectures;
- We show that iBOT is beneficial for histopathology, outperforming other in-domain pre-trained SSL networks on weakly-supervised tasks;
- We analyze the scalability of MIM through three axes: pre-training dataset size (4M^1^ to 43M patches), pre-training dataset diversity (colon-specific and pan-cancer cohorts) and architecture (22M to 307M parameters). Based on our experiments, we provide off-the-shelf guidelines on MIM pre-training with histopathology images;
- We provide slide-level features of our iBOT pre-trained models along with detailed code, weights, and documentation to reproduce our results^2^.

**Fig. 1.**
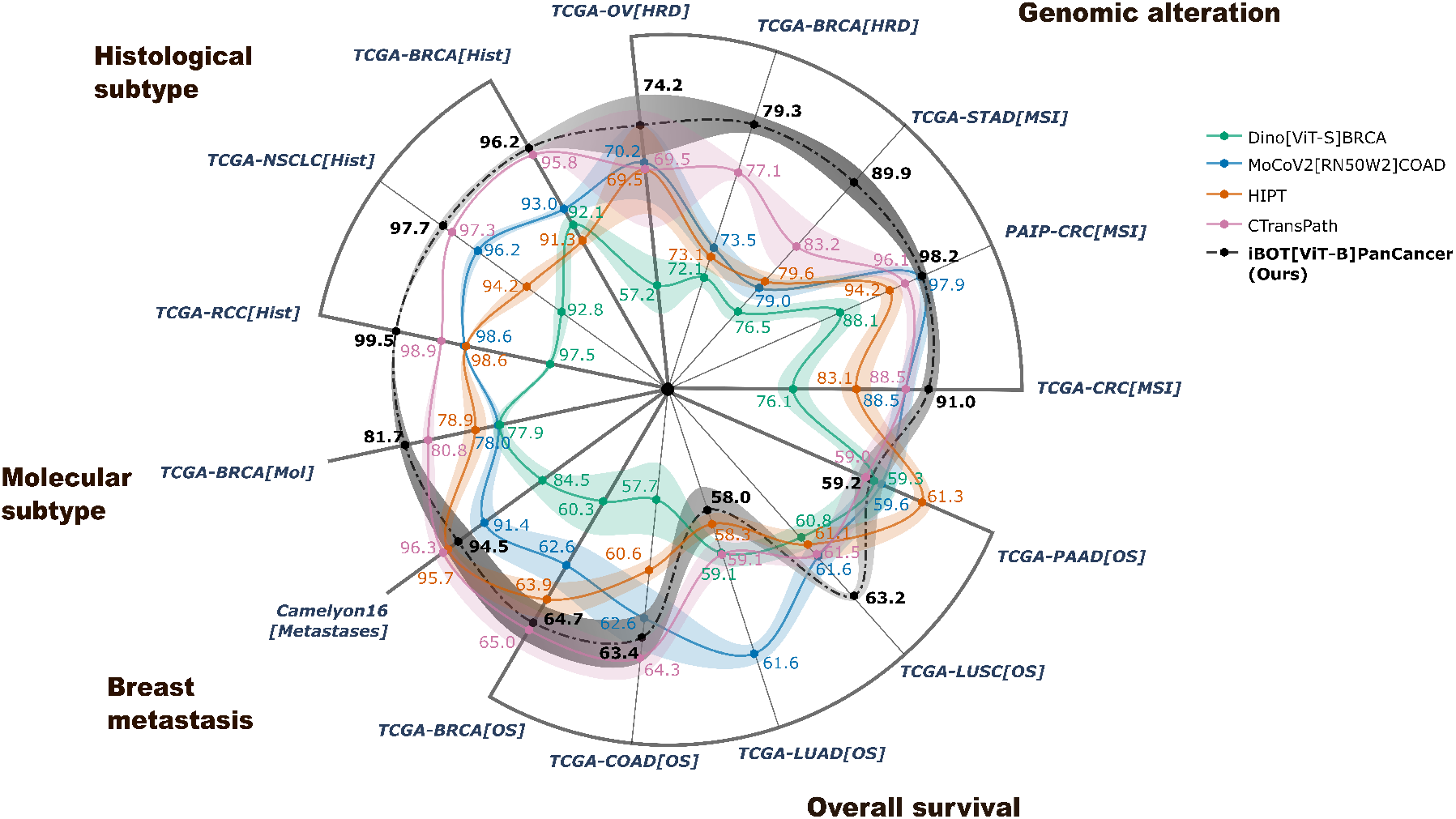
Nested cross-validation performance of our ViT-B model pre-trained with iBOT on slide-level downstream tasks against other self-supervised frameworks. We take the best performance achieved among five multiple instance learning algorithms. A 5 × 5 nested cross-validation is applied without repetition. We report the average test metrics and standard deviation on the outer folds. [MSI], [HRD], [Ctype], [Mol], [Hist] and [OS] denote respectively: MSI, HRD, Cancer Type, Molecular Subtyping, Histological Subtyping classification, and Overall Survival prediction. ROC AUC score and Harrell C-Index ([OS] suffix) are shown for classification and survival tasks, respectively. Best viewed in color.

**Fig. 2.**
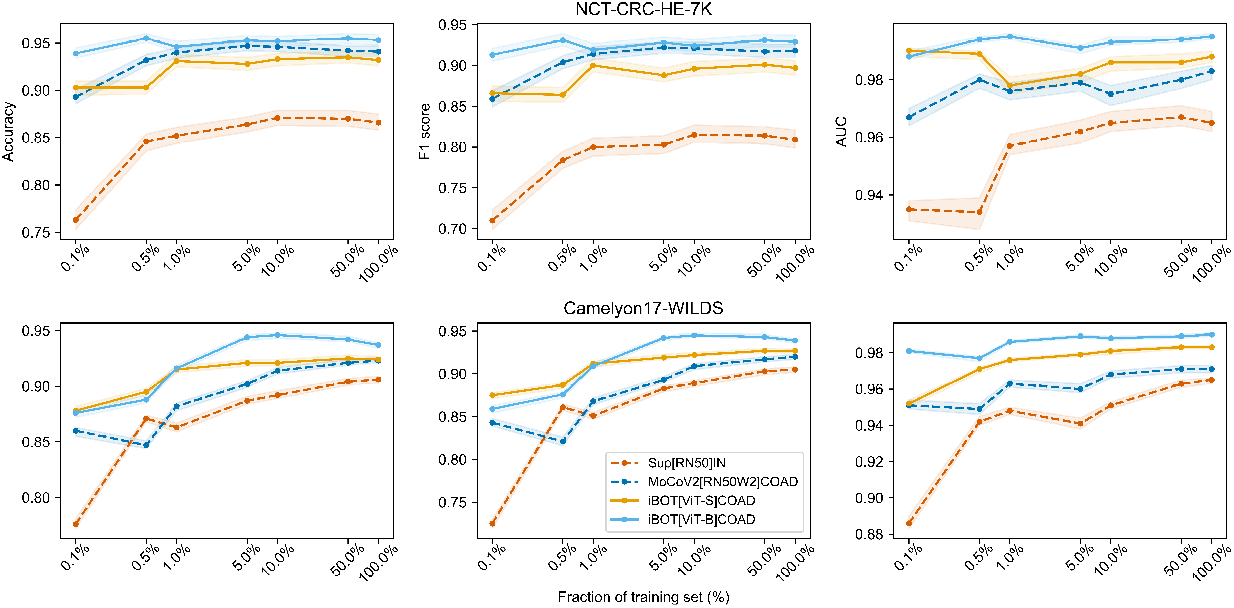
Linear evaluation results on NCT-CRC-HE and Camelyon17-WILDS testing dataset with different sizes of training data. We compare i) in-domain pre-training vs out-of-domain training for *<* 25M parameters models (dashed), ii) MoCoV2 vs iBOT methods with *>* 60M parameters models pre-trained on TCGA-COAD dataset (plain). Metrics are reported for an ensemble of 30 linear classifiers with different initializations. 95% confidence intervals are computed using bootstrap with 1,000 repeats.

## 2 Related Work

### 2.1 Self-Supervised Learning for Digital Pathology

In recent years, the field of digital pathology has remarkably benefited from advances in self-supervised learning. SSL has allowed researchers to pre-train large neural networks from massive databases of unlabeled WSI, such as The Cancer Genome Atlas (TCGA) which includes nearly 30,000 WSI from 25 anatomic sites and 32 cancer subtypes along with associated clinical, genomic, and radiomic data. In particular, CL methods such as SimCLR (36) and MoCo v2 have become quite popular. These methods rely on the idea of bringing closer, in an embedding space, pairs of similar images (*i*.*e*., positive pairs) and pushing further apart pairs of dissimilar images (*i*.*e*., negative pairs). In (17), MoCo v2 was used to pre-train a ResNet50 (37) on 2.6M patches (or tiles) from WSI of the colon adenocarcinoma cohort of TCGA (TCGA-COAD). Their experiments on Camelyon16 (breast cancer metastasis detection) showed that in-domain pre-training significantly outperforms out-of-domain (ImageNet) pre-training. The work of (38) further illustrated the benefits of in-domain pre-training. They used SimCLR to pretrain several ResNet networks on 206 thousand tiles from a total of 57 datasets and found that in-domain pre-training outperforms ImageNet pre-training on five classification and one regression tasks, while remaining comparable to ImageNet pre-training on two segmentation tasks.

While these contributions focus on CL, non-contrastive methods have also been successfully applied to digital pathology. They include Barlow Twins (39), SwAV (40) and DINO (27). While Barlow Twins shares similarities with CL methods, it does not rely on negative pairs. By bringing the cross-correlation matrix of embeddings closer to the identity matrix, Barlow Twins forces embeddings of images with similar semantic content to be closer, while penalizing redundancy among the coordinates of the embedding vectors. As opposed to MoCo v2 or Barlow Twins, SwAV takes advantage of CL methods without the need for computing pairwise comparisons. It uses online clustering and enforces consistency between cluster assignments for augmented views of the same image. SwAV learns by predicting the cluster assignment of a view given the embedding of another view (of the same image). Finally, DINO is designed to leverage the effectiveness of ViT (26). DINO uses self-distillation (with no labels): two ViT (the *teacher* and the *student*), with different parameters, compute embeddings for two augmented views of the same image, their similarity is measured using a cross-entropy loss.

In (12), the authors present a large and comprehensive study in which they investigated the impact of pre-training image encoders (CNN or ViT) on tiles from WSI images using the four SSL methods mentioned above: MoCo v2, SwAV, Barlow Twins and DINO. To this end, they extracted a total of 32.6 million tiles from TCGA cohorts and an internal dataset of WSI. The pre-trained image encoders are eventually benchmarked on four classification tasks and one instance segmentation task. The authors adapted these SSL methods to digital pathology by using color augmentations well-suited to digital pathology images. Their results show that, although no SSL method clearly outperformed the others, ViT pre-trained with DINO often provided the best performance in classification tasks. Such results encourage the use of ViT and dedicated SSL methods to efficiently pre-train these networks on large databases of WSI. Their evaluation only considers patch-level tasks and does not compare with state-of-the art frameworks tailored for histology (16, 24). In contrast, we evaluate our method against recent SSL methods available in the literature on a comprehensive list of downstream tasks both at the patch and slide-level.

### 2.2 Introduction to Masked Image Modeling

Masked Image Modeling is a recent adaptation of Masked Language Modeling in the context of computer vision. In MLM, a neural network, often based on transformers (41), is trained to predict the masked tokens in a sentence based on the context given by the non-masked tokens. This task has revolutionized the field of Natural Language Processing with the introduction of BERT (29), enabling the pre-training of very large language models on massive amounts of data (42, 43). Inspired by this work, MIM randomly masks portions of an image (patches or pixels) and learns meaningful representations by reconstructing those masked portions. The concept of MIM was first explored by (44), introducing a context encoder to mask rectangular areas and predict missing pixels. The work of (31) on Masked AutoEncoder (MAE) allowed to take advantage of ViT in the context of MIM and reinforced the interest for such methods in the context of SSL.

MLM heavily relies on the use of language tokenizers to split sentences into tokens (*e*.*g*., words, parts of a word or characters). In MIM, the design of a convenient vision tokenizer plays a crucial part as this tokenizer is used to encode the masked patches. Some studies focus on predicting the raw pixel values (32, 45) or batch-normalized pixel values (31), with the tokenizer being the identity mapping. Others use a pre-trained discrete variational autoencoder (30) or a clustering-based tokenizer that groups red-green-blue (RGB) values into k-means clusters (46), both techniques relying on a large corpus of natural images. Notably, BEiT method (30) creates discrete visual tokens using a pre-trained dVAE (47). However, the previous tokenizers often struggle to model high-frequency details and may require offline pre-training with domain-specific images. To overcome these limitations, the iBOT framework (image BERT pre-training with Online Tokenizer, (33)) proposes a single-stage pipeline where the tokenizer and the backbone encoder are jointly optimized through knowledge distillation. In addition to achieving state-of-the-art results in downstream tasks like classification, object detection, instance and semantic segmentation, iBOT exhibits robustness property against various perturbations, such as background change and occlusion. This property is particularly relevant for histopathology images, which may contain texture artifacts (blur, tissue folding, dark spots, markers or air bubbles) with potential impact on diagnostic models (25, 48). Given its high analogy with MLM, we expect MIM to improve the performance of visual models, both in terms of architecture and data scaling (31, 35, 49). The study conducted by (35) systematically explores the data scaling capability of MIM using the SimMIM method and a one billion parameters SwinV2-S model. The authors investigate the effects of different model sizes, pre-training dataset sizes, and training lengths on MIM performance. The findings suggest that, with an appropriate training length, MIM exhibits properties of being both model and data scalable.

A significant contribution of iBOT is to cast self-distillation as a token-generation self-supervised objective. An online tokenizer (the *teacher*) is fed with the original image, while the encoder (the *student*) receives a corrupted, partially masked image. The student aims to predict the correct teacher visual tokens for each masked patch token using standard Exponential Moving Average. The overall training objective of iBOT is twofold: performing self-distillation on masked patch tokens to capture low-level details and self-distillation on the class token to acquire high-level visual semantics. This novel approach eliminates the requirement for an extra pre-training phase and performs tokenization directly within the target domain.

Note that the iBOT framework is tailored for vision transformers. ViT rely on a self-attention mechanism inspired by the Transformer architecture (41), which has been highly successful in natural language processing tasks. By breaking down images into smaller patches, self-attention enables each patch to attend to all other patches, effectively modeling local information, spatial structure, and enforcing long-range dependencies in the early layers (26). Although CNN have dominated computer vision for many years, ViT have shown promising results and improved generalization, notably in histology (12, 16, 24, 50).

## 3 Pre-training setup

This section details the pre-training setup applied to our iBOT models. Sections 3.1 and 3.2 focus respectively on the different ViT architectures and pre-training datasets used in this study. In section 3.3, we provide some technical details on how the pre-training was conducted.

### 3.1 Description of ViT models used for pre-training with iBOT

To assess the scalability of the iBOT framework, we pre-trained five different models with varying architectures (ViT-S, ViT-B or ViT-L), size of the pre-training datasets (4.4M or 43.3M) and diversity (colon-specific or pan-cancer patches), as summarized on 1. Following iBOT (33), we use ViT models with different numbers of parameters: ViT-S/16 (21.7M), ViT-B/16 (85.8M) and ViT-L/16 (307M) where “/16” denotes a patch size of 16 × 16 pixels, which we omit in the next section in favor of ViT-S, ViT-B and ViT-L, respectively. Histology tiles (extracted from WSI) having a fixed size of 224 × 224 pixels, each image is represented as a grid of 14 × 14 non-overlapping tokens.

Following the DINO methodology (27), we perform multi-crop data augmentation (we refer the reader to (30) for a detailed description). For each histology tile, two global crops and ten local crops are sampled within a proportion of (32%, 100%) and (5%, 32%) of the original image size, respectively. Global and local crops are resized to 224 × 224 pixels and 96 × 96 pixels tiles, respectively. As described in (33), random MIM is performed only on the two global crops: either no cropping is applied with probability 0.5, or a proportion *p* of tokens, uniformly sampled in range [10%, 50%] of the 196 tokens, is masked out with probability 0.5. Data augmentation is performed on all crops using iBOT standard transformations: flipping, color jitter, grayscale, gaussian blur and solarization (augmentations may slightly differ between crops, see (33) and associated GitHub repository (51) for additional details).

### 3.2 Pre-training datasets

ViT models are pre-trained using iBOT on unlabeled formalin-fixed, paraffin-embedded (FFPE) hematoxylin and eosin (H&E) stained diagnostic WSI from TCGA. For each slide, non-overlapping tiles are extracted at 20 × magnification (0.5 *µ*m*/*px) with a fixed size of 224 × 224 pixels. Prior to extraction, a bi-directional U-Net neural network (52) is used to segment tissue on the input WSI and discard background and artifacts. Unless specified explicitly, a minimal tissue matter proportion of 60% is used as a selection criterion. Finally, a uniform number of tiles is sampled across slides not to exceed 4M tiles per TCGA cohort. To investigate the effect of data scaling on downstream tasks performance, we generate three datasets, denoted as TCGA-COAD, PanCancer4M and PanCancer40M. TCGA-COAD pre-training dataset contains a total of 441 slides and 4,386,755 tiles for 434 patients with colon adenocarcinoma. PanCancer40M pre-training dataset covers 13 anatomic sites and 16 cancer subtypes for 5,558 patients, representing a total of 6,093 slides and 43,374,634 patches. Finally, PanCancer4M is a subset of PanCancer40M with 5,183WSI and 4,386,755 tiles overall (see Table B1 and Table B2 in appendix for cohorts distribution).

### 3.3 Pre-training details

To ensure stability in the pre-training stage, we pre-trained our ViT models with iBOT using specific sets of parameters, depending on the size of the architecture. This section gives an overview of the most influential parameters, further details can be found in appendix.

Teacher temperature was set to 0.04 with an initial value of 0.04 and 30 warm-up epochs for iBOT ViT-B and iBOT ViT-L, ten warm-up epochs for iBOT ViT-S. AdamW optimizer (53) was used and learning rate linearly ramped up during the first ten (resp. three) epochs for ViT-B and ViT-L (resp. ViT-S) to its base value scaled with the total batch size according to: 0.0005 × *B/*256 (54), where *B* denotes the batch size. The final learning rate was set to 0.000002 through a cosine schedule. Regarding multi-crop augmentation, two global crops and ten local crops are sampled within a proportion of (32%, 100%) and (5%, 32%) instead of (14%, 100%) and (5%, 40%), respectively.

All models were implemented in PyTorch 1.13.1 and pre-trained on 16 to 64 NVIDIA V100 GPUs with 32Gb RAM on the French Jean Zay cluster. Batch size and corresponding time to convergence are reported in appendix (Table B1) for each iBOT model.

## 4 Experimental and evaluation setup

After pre-training, we evaluate our ViT models pre-trained with iBOT on a wide range of downstream tasks. This includes 17 datasets covering seven cancer indications. Slide-level experiments involve 14 weakly-supervised WSI classification tasks using TCGA cohorts, Camelyon16 and PAIP-CRC datasets. Patch-level experiments are conducted on the NCT-CRC-HE and Camelyon17-WILDS (55) datasets with two patch classification tasks. Detailed information about the datasets, tasks and validation protocol used in our experiments is provided in this section.

### 4.1 Slide-level experiments

The first category of downstream experiments consists of slide-level prediction tasks on a various range of outcomes (histological and molecular subtypes, genetic alterations, cancer types, overall survival). We describe them in this section.

#### 4.1.1 Downstream tasks and corresponding datasets

##### Histological subtype prediction TCGA-RCC

Renal cell cancer (RCC) can be divided into three histological subtypes. The goal of this classification task is to classify each slide as: kidney renal clear cell carcinoma (KIRC), kidney renal papillary cell carcinoma (KIRP) or kidney chromophobe (KICH). **TCGA-BRCA**. Breast carcinoma (BRCA) can be divided into two main histological subtypes. The goal of this task is to classify each slide as either invasive lobular carcinoma (ILC) or invasive ductal carcinoma (IDC).

##### Molecular subtype prediction TCGA-BRCA

This multi-class classification task aims at distinguishing between normal-like (Normal), basal-like (Basal), luminal A (LumA), luminal B (LumB) and Her2-enriched (Her2) molecular subtypes.

##### Cancer type prediction TCGA-NSCLC

Non-small cell lung carcinoma (NSCLC) is commonly divided into two main cancer types. For a given slide, this classification task aims at predicting the lung adenocarcinoma (LUAD) or lung squamous cell carcinoma (LUSC) cancer type.

##### Genomic alterations prediction MSI and HRD

MSI and HRD are both abnormalities impacting the deoxyribonucleic acid damage repair (DDR) process in tumors. Early recognition of those biomarkers may benefit the patients through specific therapies targeting DDR-related genomic alterations (19, 20). This is of particular interest in breast cancer (21, 56) and colorectal cancer (11, 21). For MSI or HRD, we aim at predicting high vs low instability or deficiency (MSI-H vs MSS/MSI-L, or HRD-H vs HRD-L) patients. **MSI on PAIP-CRC**. PAIP (57, 58) provides 2,547 WSI collected from three Korean centers (Seoul National University Hospital, Seoul National University Bundang Hospital and SMG-SNU Boramae Medical Center), covering six cancer types. We retrieved 47 patients from PAIP with colorectal tumors and available MSS/MSI-L labels, provided by the Pathology AI Platform. This dataset is used for external validation only after training on the TCGA-CRC cohort.

##### Metastases detection Camelyon16

Camelyon16 (59) is a dataset of H&E stained slides from lymph node sections designed for the automated detection of metastasis in breast cancer. This dataset contains 399 WSI from two medical centers, divided into 269 training and 130 testing WSI. In this work, we consider slide-level labels indicating whether a given WSI contains metastases or not.

##### Overall survival OS

The OS refers to durations between the beginning of treatment and potential all-cause mortality. OS prediction task aims to estimate time-to-events, taking right-censoring into account, *i*.*e*., potential loss of follow-up or no event before the end of the study.

#### 4.1.2 Models: weakly-supervised learning

The weakly-supervised classification problem in whole slide images involves providing global annotations at the slide level without detailed pixel-level annotations for internal regions. Existing weakly-supervised algorithms for WSI classification typically consist of two main steps: patch-level feature extraction within the WSI and subsequent feature aggregation using multiple instance learning (MIL) algorithms. To evaluate the intrinsic representation capacity of the different SSL models, we highlight the results obtained with two MIL algorithms, namely ABMIL (60) and TransMIL (61). These algorithms are applied to features extracted from WSI patches. To further illustrate the impact of the choice of MIL model, some of our results also report results obtained with the following MIL algorithms: DSMIL (62), Chowder (10) and MeanPool, the latter simply averaging patch features across the slide to obtain a single slide-level representation, and applying a multi-layer perceptron (MLP) on it. In particular, note that the comparison in section 5.3 highlights the optimal performance across those five MIL algorithms. For a comprehensive review of aggregation methods in weakly-supervised learning, including MIL, see the recent work of (63).

During the training process, we retain a random subset of 1,000 tile features for each slide. The Adam optimizer is utilized with a mini-batch size of eight slides. The MIL algorithms are trained for a maximum of 50 epochs for SSL in-domain models, and 100 epochs when utilizing a ResNet-50 pre-trained on ImageNet. This additional training time is considered to account for the out-of-domain pre-training task (*i*.*e*., classification of object-centric natural images), which may lead to less linearly separable features of histology images, hence to a more unstable training.

For all tasks, we optimize binary cross-entropy loss for binary classification tasks and categorical cross-entropy loss for multi-class classification tasks. To estimate overall survival, we employ a differentiable Cox loss (64, 65). cross-validation (CV) folds are created at the patient level and stratified based on class distribution or censoring proportion.

#### 4.1.3 Evaluation with nested cross-validation

In a real-case machine learning scenario, one has to simultaneously select the best model for a given dataset and assess its generalization performance. Even though model selection is different from model generalization assessment (66), most works report the cross-validation error found for the optimal model during the model selection as the assessed model generalization performance (67). (67–69) report a bias in error estimation when using cross-validation for model selection and model assessment simultaneously. Indeed, hyperparameter optimization can lead to overfitting a dataset or a specific data split, and provide an over-optimistic evaluation of the actual model performance, that should not be used for model generalization evaluation. (67, 69) suggest to rather use nested cross-validation (or double cross-validation) as an unbiased estimate of the true error. Nested cross-validation involves two levels of cross-validation, an outer and inner cross-validation. Within the training outer folds, an inner cross-validation is performed for hyperparameter tuning and model selection. The best model configuration is chosen based on the average performance across the inner folds. This selected model is then evaluated on the corresponding validation outer fold, which was not used during model selection. The performance metrics obtained from each validation outer fold are averaged to estimate the model generalization performance. This eliminates the bias introduced by standard cross-validation procedure as the test data in each iteration of the outer cross-validation has not been used to optimize the performance of the model in any way, and may therefore provide a more reliable criterion for choosing the best model.

In this study, we applied stratified nested cross-validation to reduce the bias of the resulting error estimate. As such, we perform model selection (hyperparameter tuning) and model assessment through 5×5 nested cross-validation with no repeats (five inner and five outer splits). During nested-CV, we test different values of the initial learning rate and weight decay, namely *{*0.001, 0.0001*}* for learning rate and*{* 0, 0.0001 *}* for weight decay, respectively. The optimal number of epochs is determined within each outer split through the 5-fold inner CV based on the validation metric. One of the tasks includes the evaluation on an external cohort using the PAIP-CRC cohort for MSI prediction, with TCGA-CRC serving as the training set. In this scenario, we employed standard 5-fold CV with three repeats on the internal training cohort TCGA-CRC. This allows us to create an ensemble of 15 models that is subsequently evaluated on the PAIP-CRC external cohort. Hyperparameter tuning is conducted on the internal training cohort using the same set of configurations as in the nested CV approach.

### 4.2 Patch-level experiments

The second category of downstream experiments consists of patch-level classification tasks, which we described below.

#### Downstream tasks and corresponding datasets

##### Colorectal tissue phenotyping

In the NCT-CRC-HE (70) datasets, the task consists in classifying each patch of colorectal cancer image as one of nine tissue types: Adipose (ADI), background (BACK), debris (DEB), lymphocytes (LYM), mucus (MUC), smooth muscle (MUS), normal colon mucosa (NORM), cancer-associated stroma (STR) and colorectal adenocarcinoma epithelium (TUM). The training set, NCT-CRC-HE-100K, consists of 100,000 patches extracted from 86 WSI at a resolution of 0.5 *µm/*px, collected at the NCT Biobank National Center for Tumor Diseases (Heidelberg, Germany) and the UMM pathology archive (University Medical Center Mannheim, Mannheim, Germany). All patches have a size of 224 × 224 pixels. NCT-CRC-HE-7K serves as an independent set of 7,180 patches used for testing. Images without Macenko (71) normalization were used.

##### Metastases detection

Camelyon17-WILDS (55, 72) dataset is a patch-based variant of the Camelyon17 (73) dataset. It contains 450,000 H&E stained lymph-node scans from five hospitals, extracted at 20× magnification (0.5 *µm/*px resolution) from 50 WSI. The binary classification task aims at detecting the presence of metastasis on 96 × 96 pixels breast cancer patches from lymph nodes sections, in the presence of high domain shifts between hospitals. The training set is composed of three centers (ID 0, 3 and 4) for a total of 335,996 patches, whereas the testing set only contains center 1 with 34,904 patches, considered as out-of-distribution.

#### 4.2.1 Linear evaluation

Patch classification tasks are evaluated on top of pre-extracted frozen features. Patches are resized according to the input shape of the ViT or CNN (224 × 224 pixels or 256 × 256 pixels). Linear evaluation is evaluated by training a logistic regression with stochastic gradient descent optimization for 1,000 iterations. Early stopping is performed on 10% of training data, serving as a validation set. The initial value of the learning rate is set to 0.0001 and is divided by five each time the validation loss does not decrease for five consecutive epochs. We perform an ensemble of 30 different models with different random initialization and data shuffling. Linear evaluation is performed at different sizes of the training datasets, namely 0.1%, 0.5%, 1%, 5%, 10%, 50% and 100%.

### 4.3 Other representation learning frameworks

To further demonstrate the validity of our iBOT-pretrained ViT models, we conduct a comprehensive comparison with existing other representation learning methods. Those feature extraction methods include ImageNet-pretraining and SSL-pretraining, described as follows:

- CTransPath (24): a hybrid model composed of a CNN and a multi-scale Swin transformer architecture. CTransPath implements SCRL, a semantically-relevant contrastive learning method, an extension of MoCo v3 (74) method implementing an additional branch where positive pairs no longer come from the same instance, but rather a large memory bank of pseudo-positives, semantically-relevant images. CTransPath was pre-trained on 14.3M unlabeled patches at 20× magnification with a size of 1024 × 1024 pixels.
- HIPT (16): a hierarchical image pyramid ViT trained with DINO (27). A first patch-level ViT is trained on 256 × 256 pixels images (20 ×). From the output tokens of the first ViT-S, 4096 × 4096 pixels images are encoded into 16 × 16 pixels tokens, on which a second region-level ViT-Tiny (2.8M parameters with output dimension 182) is trained to produce region-level tokens. Patch and region-level being frozen, tokens from 4096 × 4096 pixels regions from a given WSI are finally aggregated using a final transformer fine-tuned on downstream tasks. Pre-training dataset covers 33 cancer types from TCGA, from which patch-level and region-level ViT were trained on 104M patches and 408,218 regions respectively.
- Dino[ViT-S]BRCA (formalism detailed below): (50) pre-trained a ViT-Small model with DINO. The pre-training dataset consists of two million patches with shape 256 × 256 pixels, extracted from 1,038 WSI in the TCGA-BRCA cohort.
- MoCov2[RN50W2]COAD (formalism detailed below): (17) pre-trained a wide ResNet-50-2 (75) with MoCo v2 on the TCGA-COAD cohort. Tiles were extracted at 20× magnification with a fixed shape of 224 × 224 pixels. Those tiles are identical to that of the TCGA-COAD dataset described in section 3.2.

As opposed to the previous in-domain SSL methods, we also consider one out-of-domain supervised method to further compare the impact of pre-training domains on downstream tasks performance. We use a ResNet-50 (37) pre-trained on Imagenet-1K (1.2M natural images).

Weights from DinoBRCA, HIPT and CTransPath models were retrieved directly from their respective GitHub repositories (see appendix C for details). MoCoV2COAD was pre-trained from scratch on the TCGA-COAD dataset, following the same training recipe and hyperparameters settings as the original publication (See table 1).

**Table 1.** Description of ViT models pre-trained with iBOT on histology tiles for this study. We provide a description of MoCoV2[RN50w2]COAD for comparison with a purely contrastive learning framework. Model name formalism is described at the end of section 4.3. GPU: Graphical Processing Unit.

| Model name | No. params | Size of pre-trained datasets | Batch size per GPU | No. V100 GPUs | Total batch size | No. iter | Training time (GPU hours) |
| --- | --- | --- | --- | --- | --- | --- | --- |
| MoCoV2[RN50w2]COAD | 66.8M | 4.4M | 256 | 16 | 4,096 | 215k | 2,300 |
| iBOT[ViT-S]COAD | 21.7M | 4.4M | 112 | 16 | 1,792 | 245k | 1,152 |
| iBOT[ViT-B]COAD | 85.8M | 4.4M | 60 | 24 | 1,440 | 165k | 1,704 |
| iBOT[ViT-L]COAD | 307M | 4.4M | 20 | 64 | 1,280 | 165k | 3,712 |
| iBOT[ViT-S]PANCAN | 21.7M | 4.4M | 112 | 16 | 1,792 | 245k | 1,152 |
| iBOT[ViT-B]PANCAN | 86M | 43.3M | 45 | 32 | 1,440 | 155k | 1,216 |

In the following sections, models are named using the <monospace>framework[architecture]pre-training-dataset </monospace>formalism. Accordingly, the following model names denote:

- *MoCoV2[RN50W2]COAD*: a ResNet-50-w2 pre-trained with MoCo v2 SSL framework on TCGA-COAD dataset
- *Sup[RN50]IN*: a ResNet-50 pre-trained in a supervised fashion on ImageNet-1K.
- *iBOT[ViT-X]COAD*: a ViT-S, ViT-B or ViT-L pre-trained with iBOT SSL framework on TCGA-COAD
- *iBOT[ViT-S]PanCancer*: a ViT-S pre-trained with iBOT on TCGA-COAD dataset
- *iBOT[ViT-B]PanCancer*: a ViT-B pre-trained with iBOT on PanCancer40M dataset
- *Dino[ViT-S]BRCA*: a ViT-S pre-trained with DINO on TCGA-BRCA dataset

### 4.4 Metrics

Slide-level prediction tasks performance is evaluated using the area under the receiver operating characteristic curve (ROC AUC), while Harrell C-index (76) is utilized for OS prediction tasks. Standard deviations are consistently reported for nested CV and calculated across the five outer folds. We use bootstrap on 1,000 repeats with replacement to generate a 95% confidence interval for ROC AUC scores obtained with PAIP-CRC external validation. Bootstrap hypothesis testing is used to statistically compare the mean performance of our models on PAIP-CRC external cohort. Patch-classification tasks performance is evaluated using the accuracy, F1 and ROC AUC score. Macro F1 and ROC AUC are reported for multi-class classification tasks. It should be noted that the “background” (BACK) class is not considered for NCT-CRC-HE neither for training nor for evaluation, following (23, 50, 77, 78).

## 5 Results

In this section, we first provide a comparison of iBOT pre-trained ViT models against out-of-domain SSL pre-trained models and a purely contrastive model pre-trained from scratch on TCGA-COAD using MoCo v2. Then, we investigate the effect of scaling iBOT models in terms of architecture (ViT-S to ViT-L), dataset size and diversity (TCGA-COAD vs. PanCancer 40M). Finally, we compare our iBOT[ViT-B]PanCancer to state-of-the-art in-domain architectures.

As described in section 4, we report results on both weakly-supervised WSI classification and patch-level classification tasks:

- For each WSI-level task, slide features are generated from a given model, which may be trained in a supervised fashion (ResNet50 on ImageNet) or pre-trained on in-domain pathology datasets (see section 4.3). Then, feature aggregation is performed through a MIL algorithm, which is trained and optimized using nested CV. In the following sections, unless specified otherwise, results for nested CV or external validation are depicted for two MIL algorithms: ABMIL and TransMIL. All results are re-produced using the original implementation and corresponding released codes.
- Patch-level classification tasks are assessed using a linear evaluation protocol described in section 4.2.1. We conduct experiments on training with different fractions to investigate the classification performance under limited labeling, by randomly sampling 0.1% to 100% of the training data.

### 5.1 In-domain pre-training with iBOT

#### 5.1.1 Comparison of in-domain and out-of-domain pre-training

In this section, we discuss the advantages of using in-domain pre-trained neural networks compared to out-of-domain pre-trained ones, namely models pre-trained on the ImageNet database. In Table 2, the first (*i*.*e*., Sup[RN50]IN) and second (*i*.*e*., iBOT[ViT-S]COAD) columns show that using a feature extractor pre-trained on patches from WSI brings a consistent improvement over a vast majority of downstream tasks. We observe an improvement of up to 4.0%, on average, across all downstream tasks using ABMIL (both in terms of ROC AUC for classification tasks and Harrell’s C-Index for survival analysis tasks). This average improvement increases by up to 7.9% on colorectal cancer-specific tasks, namely MSI prediction, with a remarkable generalization performance on PAIP-CRC external validation (92.1 vs. 78.7 ROC AUC on MSI prediction, see Table 3). It is worth noting that, although the two models have different architectures, they both share a comparable number of trainable parameters (21.7M for ViT-S and 25M for ResNet50). The same conclusion can be drawn from Table 3 (external validation of the PAIP-CRC dataset). The advantage of in-domain pre-training over out-of-domain pre-training is also noticeable on patch-level classification tasks (Table 4). Indeed, statistically significant improvements (*p <* 0.0001) are observed from using in-domain pre-trained methods over Sup[RN50]IN, *i*.*e*. 6.8 (resp. 1.8), 7.5 (resp. 1.7) and 8.7 (resp. 2.9) accuracy gains on the NCT-CRC-HE-7K (resp. Camelyon17-WILDS) dataset offered by iBOT[ViT-S]COAD, MoCoV2[RN50W2]COAD and iBOT[ViT-B]COAD models. Results from Figure 3 and Table 4 also highlight the high-label efficiency and discriminative capacity of iBOT pre-trained ViT models both in very low data regime for the downstream task (*<* 5%) and full training dataset setting.

**Table 2.**
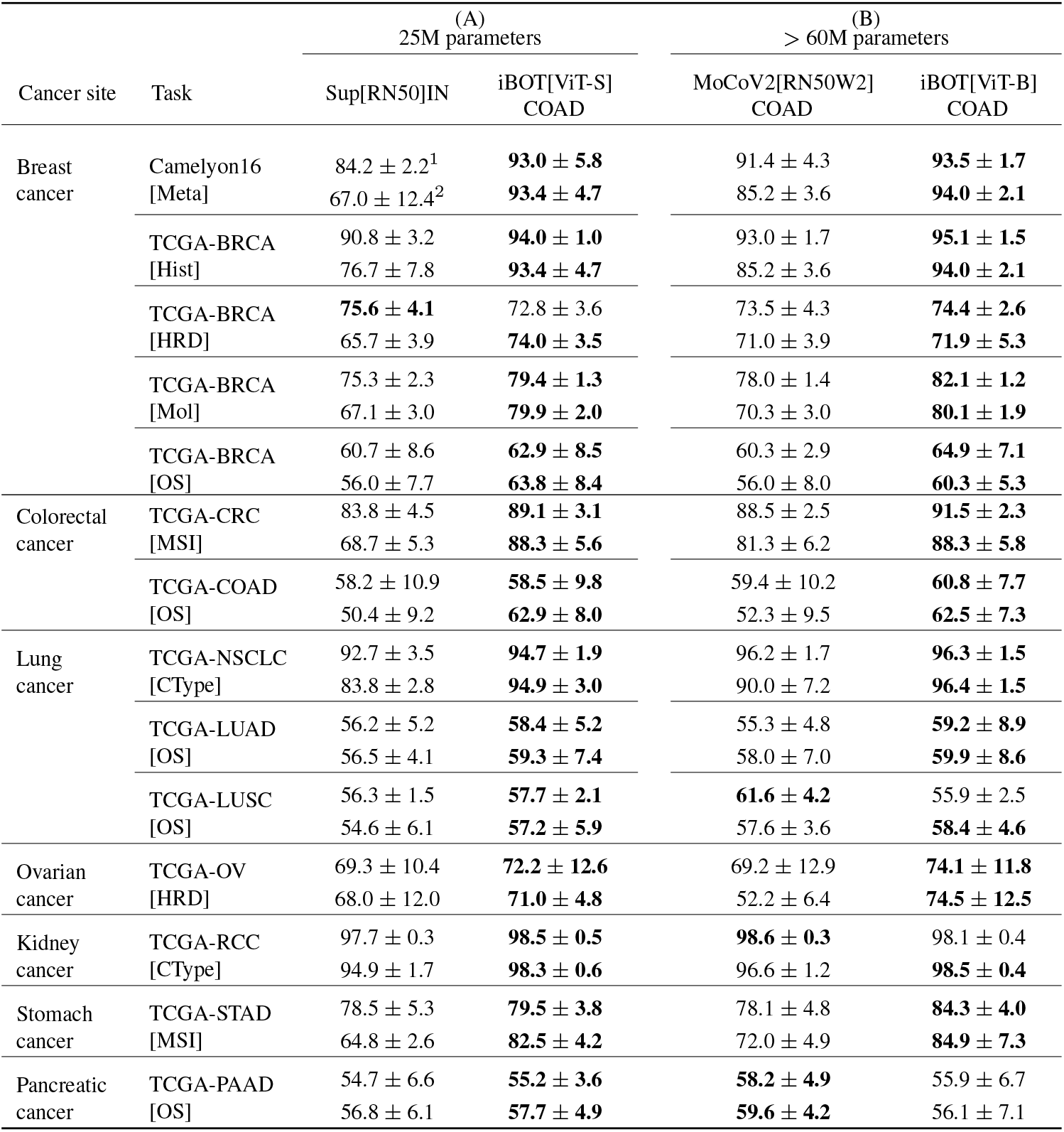
Comparison weakly-supervised downstream performance with (A) in-domain pre-training vs out-of-domain training, (B) MoCoV2 vs iBOT methods with TCGA-COAD pre-training. [MSI], [HRD], [Ctype], [Mol], [Hist] and [OS] denote respectively: MSI, HRD, Cancer Type, Molecular Subtyping, Histological Subtyping classification, and Overall Survival prediction. We take the average and standard deviation of each metric over the five outer test splits from nested CV. Bold indicates the highest performance for each MIL model in (A) and (B) separately (^1^ABMIL, ^2^TransMIL).

**Table 3.**
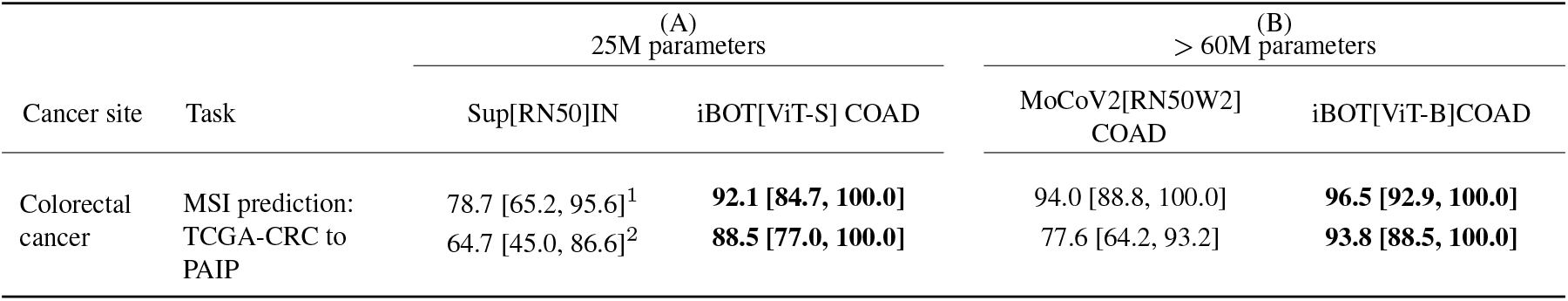
Comparison of external validation performance for (A) in-domain pre-training vs out-of-domain training, (B) MoCoV2 vs iBOT methods with TCGA-COAD pre-training. Results are reported for PAIP-CRC[MSI] external validation after training on TCGA-CRC[MSI] classification task. ROC AUC scores and 95% confidence intervals are computed using bootstrap with 1,000 repeats. Bold indicates the highest performance for each MIL model in (A) and (B) separately (^1^ABMIL, ^2^TransMIL).

**Table 4.**
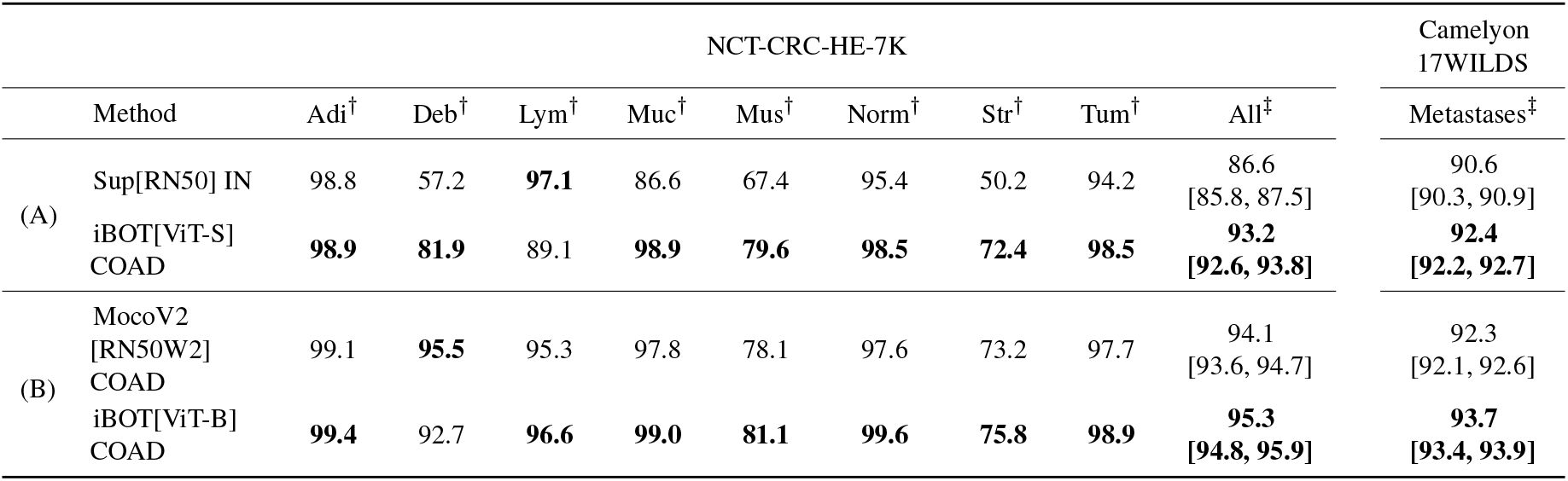
Comparison of patch classification performance for (A) in-domain pretraining vs out-of-domain training, (B) MoCoV2 vs iBOT methods with TCGA-COAD pre-training. F1 score () is reported for single class classification (ADI to TUM) in NCT-CRC-HE-7K. Accuracy (‡) and 95% confidence intervals are computed using bootstrap with 1,000 repeats for multi-class classification in NCT-CRC-HE-7K and binary classification in Camelyon17-WILDS, respectively. Bold indicates the highest performance across classes. ROC AUC scores are reported in appendix (Table G1).

| NCT-CRC-HE-7K |  |  |  |  |  |  |  |  |  | Camelyon<br>17WILDS |
| --- | --- | --- | --- | --- | --- | --- | --- | --- | --- | --- |
| Method | Adi† | Deb† | Lym† | Muc† | Mus† | Norm† | Str† | Tum† | All‡ | Metastases‡ |
| (A) Sup[RN50] IN | 98.8 | 57.2 | **97.1** | 86.6 | 67.4 | 95.4 | 50.2 | 94.2 | 86.6<br>[85.8, 87.5] | 90.6<br>[90.3, 90.9] |
| iBOT[ViT-S]<br>COAD | **98.9** | **81.9** | 89.1 | **98.9** | **79.6** | **98.5** | **72.4** | **98.5** | **93.2**<br>[92.6, 93.8] | **92.4**<br>[92.2, 92.7] |
| (B) MocoV2<br>[RN50W2]<br>COAD | 99.1 | **95.5** | 95.3 | 97.8 | 78.1 | 97.6 | 73.2 | 97.7 | 94.1<br>[93.6, 94.7] | 92.3<br>[92.1, 92.6] |
| iBOT[ViT-B]<br>COAD | **99.4** | 92.7 | **96.6** | **99.0** | **81.1** | **99.6** | **75.8** | **98.9** | **95.3**<br>[94.8, 95.9] | **93.7**<br>[93.4, 93.9] |

**Fig. 3.**
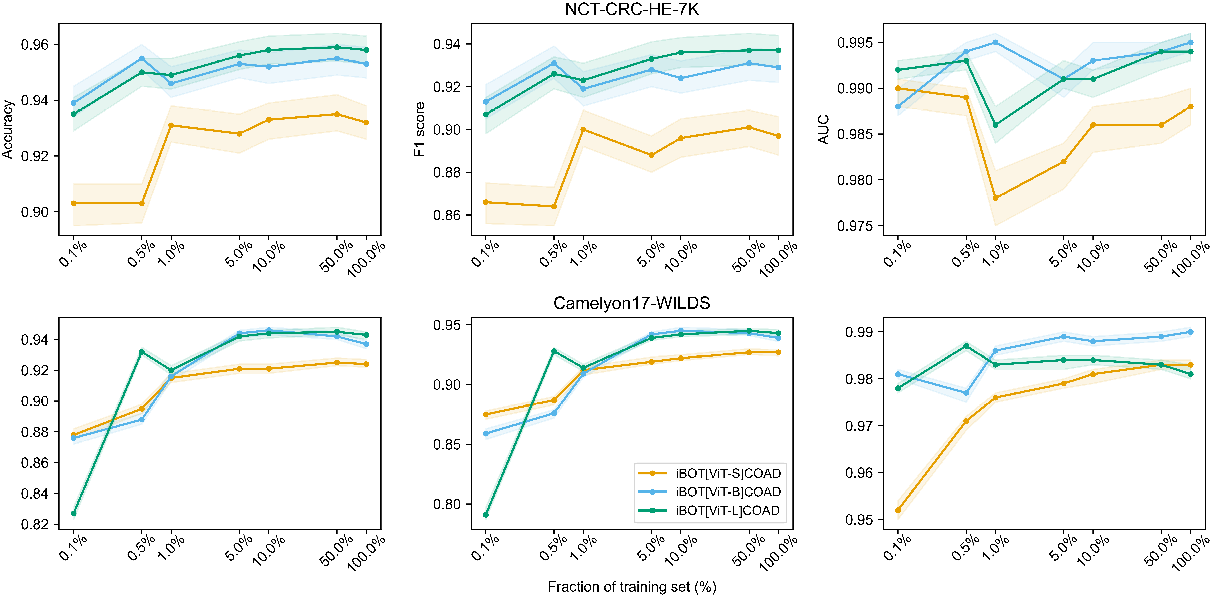
Linear evaluation results on NCT-CRC-HE and Camelyon17-WILDS testing dataset with different sizes of training data and sizes of ViT architectures. Metrics are reported for an ensemble of 30 linear classifiers with different initializations. 95% confidence intervals are computed using bootstrap with 1,000 repeats.

More specifically, we observe on NCT-CRC-HE-7K a performance plateau for all models from 10% of the training dataset while the gap between Sup[RN50]IN and other SSL methods remains constant. This suggests that the out-of-domain Sup[RN50]IN can not benefit from more examples during training. In contrast, other pre-trained models on TCGA-COAD show highly discriminative capacity on in-domain data from 0.1%, indicating that in-domain pre-training is much more beneficial to this particular downstream task. This observation is mitigated by the results on Camelyon17-WILDS, which can be seen as different degrees of an out-of-domain dataset (breast instead of colon or natural images). In this task, Sup[RN50]IN still underperforms but closes the gap in higher ratio of the training data.

These results are aligned with the conclusions from previous work (12, 38, 50). Although transfer learning from ImageNet usually provides a strong baseline, one might expect in-domain pre-training to provide consequential performance gains.

#### 5.1.2 Advantages of using iBOT for pre-training with respect to MoCo v2

In the previous section, we presented results in favor of in-domain pre-training with MoCo v2 or iBOT. A question remains: should one of these two SSL methods be preferred over the other? In this section, we argue that iBOT should be preferred to MoCo v2 for in-domain pre-training and provide experimental results on a variety of downstream tasks to support this. The second (*i*.*e*., iBOT[ViT-S]COAD) and third (*i*.*e*., MoCoV2[RN50W2]COAD) columns of Table 2 show that, despite an approximately three times smaller model and pre-trained on the same in-domain TCGA-COAD dataset, iBOT[ViT-S]COAD brings an average 0.72% performance gain over MoCoV2[RN50W2]COAD, to mitigate with an average *−* 0.93% drop across CRC-related tasks. Additionally, we also provide a comparison between MoCoV2[RN50W2]COAD (third column of Table 2) and iBOT[ViT-B]COAD (fourth column of Table 2) on multiple downstream tasks, two models pre-trained on TCGA-COAD and with roughly comparable number of trainable parameters (66.8M for Wide ResNet50-2 and 85.8M for ViT-B). Overall, iBOT[ViT-B]COAD brings a +3.2% mean improvement on MoCoV2[RN50W2]COAD across all tasks, outperforming the CL based method both on non-CRC-related (+3.3%) tasks and CRC-related (+2.7%) with a 96.5 vs. 94.0 ROC AUC score on PAIP-CRC external validation (Table 3). From the results in Figure 3 and Table 4, we also see that iBOT[ViT-B]COAD brings a strong improvement in patch-level classification tasks associated with NCT-CRC-HE-7K and Camelyon17-WILDS.

It should be noted that despite using the same validation scheme and hyperparameter tuning, the results of TransMIL for Sup[RN50]IN and MoCoV2[RN50W2]COAD do not match those of ABMIL by a large margin. This discrepancy could be attributed to the higher output feature dimension of these models (2048) which is approximately 5 (resp. 2.5) times higher than that of iBOT[ViT-S]COAD (resp. iBOT[ViT-B]COAD). We encountered difficulties with overfitting and convergence when applying TransMIL on top of 2048-dimensional features.

### 5.2 Scaling iBOT with model architecture

In this section, we examine the impact of increasing the size of the ViT model from a ViT-S (21.7M) to a ViT-L (307M) architecture. We pre-trained all iBOT models using the TCGA-COAD pre-training dataset. The results in Table 5 and Table 6 indicate that scaling the model from a ViT-S (21.7M) to ViT-B (85.8M) architecture strongly affects the performance on downstream tasks using ABMIL, with an average gain of 2.5% across all tasks. However, the results also demonstrate that further increasing the size of the architecture from ViT-B to ViT-L (307M) does not provide clear benefits. The ViT-L model leads to an overall performance loss of 0.2% and a 1.7% loss on the four OS prediction tasks compared to the ViT-B counterpart. This suggests that our ViT models reach their discriminative capacity saturation when architecture is not scaled alongside the pre-training dataset size.

**Table 5.**
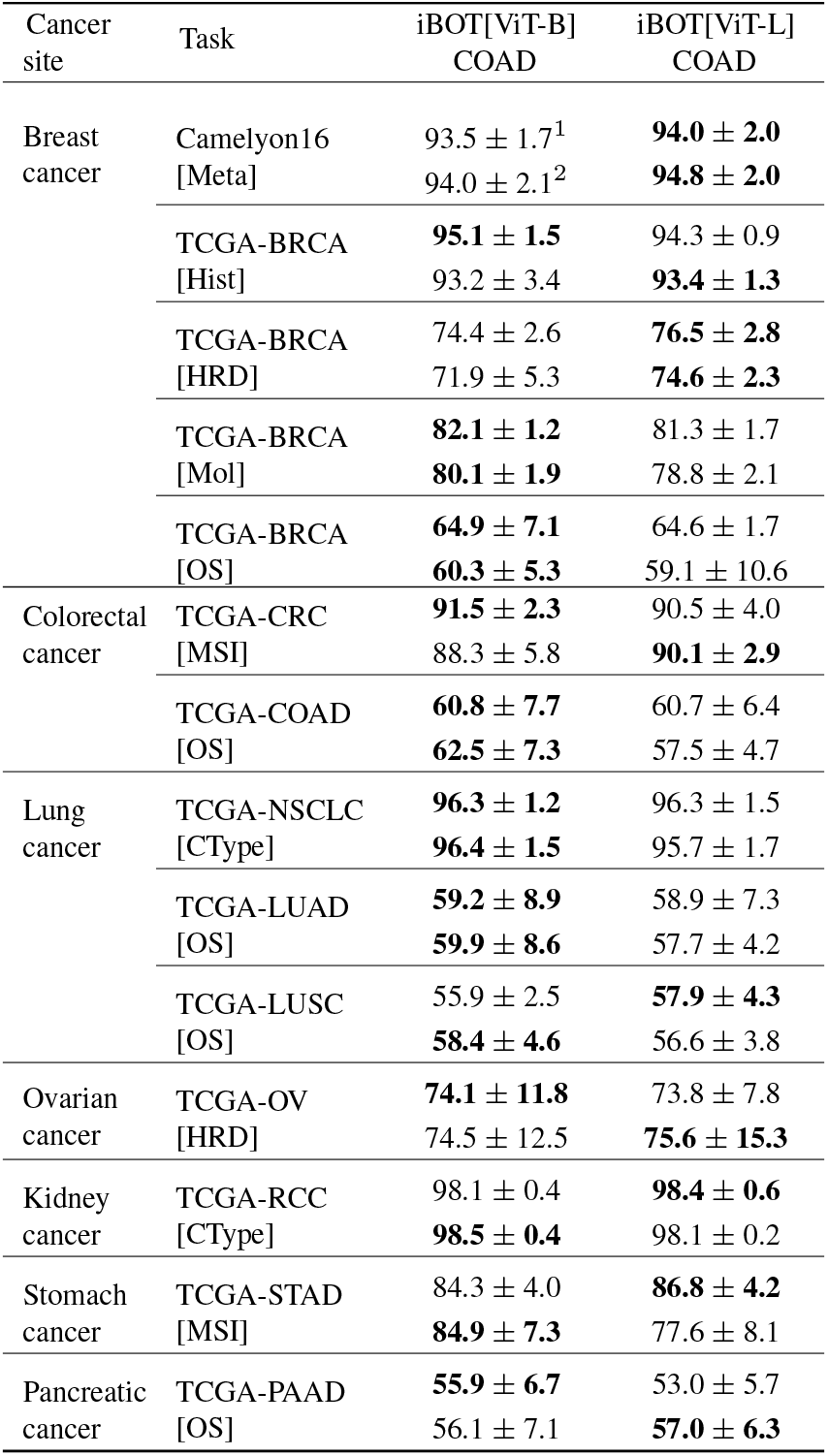
Impact of ViT architecture on weakly-supervised downstream performance. The iBOT[ViT-B]COAD column is repeated from Table 2 to ease the comparison with iBOT[ViT-L]COAD. ROC AUC scores and C-Index are reported for classification and survival tasks, respectively. We take the average and standard deviation of each metric over the five outer test splits from nested CV. Bold indicates the highest performance for each MIL model (^1^ABMIL, ^2^TransMIL).

**Table 6.**
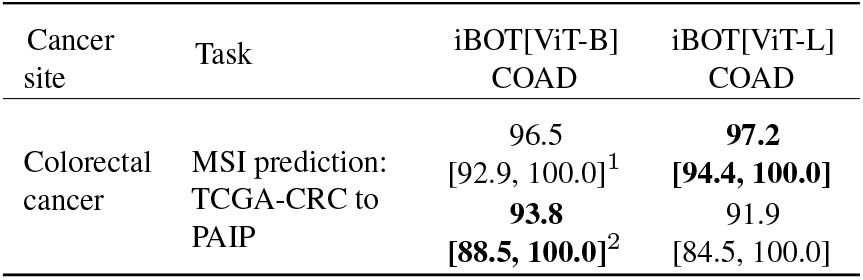
Impact of ViT architecture scaling on external validation. The iBOT[ViT-B]COAD column is repeated from Table 3 to ease the comparison with iBOT[ViT-L]COAD. ROC AUC scores and 95% confidence intervals are computed using bootstrap with 1,000 repeats. Bold indicates the highest performance for each MIL model (^1^ ABMIL, ^2^ TransMIL).

Table 7 presents the ROC AUC scores on the NCT-CRC-HE-7K and Camelyon17-WILDS test datasets. We fit a logistic regression for 100 iterations on top of the frozen patch features using 100% of the corresponding training sets. We observe statistically significant improvements (*p <* 0.0001) from ViT-S to ViT-B architectures on both datasets. Interestingly, these architectures produce similar results (accuracy, F1, and ROC AUC scores) with less than 5% of the training set on NCT-CRC-HE-7K (see Figure 3). However, in this scenario of low data regime, the ViT-B architecture performs significantly better (*p <* 0.0001) than its ViT-S counterpart on the breast cancer patches classification task, Camelyon17-WILDS. This observation suggests that smaller architectures struggle to transfer knowledge to out-of-domain tasks compared to larger ones.

**Table 7.** Impact of ViT architecture scaling on patch classification tasks. The iBOT[ViT-B]COAD line is repeated from Table 4 to ease the comparison with iBOT[ViT-L]COAD. F1 score (†) is reported for single class classification (Adi to Tum) in NCT-CRC-HE-7K. Accuracy (‡) and 95% confidence intervals are computed using bootstrap with 1,000 repeats for multi-class classification in NCT-CRC-HE-7K and binary classification in Camelyon17-WILDS, respectively. Bold indicates the highest performance across classes. ROC AUC scores are reported in appendix (Table G2).

| Method | NCT-CRC-HE-7K |  |  |  |  |  |  |  |  | Camelyon<br>17WILDS |
| --- | --- | --- | --- | --- | --- | --- | --- | --- | --- | --- |
| | Adi† | Deb† | Lym† | Muc† | Mus† | Norm† | Str† | Tum† | All‡ | Metastases‡ |
| iBOT[ViT-B]<br>COAD | **99.4** | 92.7 | 96.6 | 99.0 | 81.1 | **99.6** | 75.8 | **98.9** | 95.3<br>[94.8, 95.9] | 93.7<br>[93.4, 93.9] |
| iBOT[ViT-L]<br>COAD | 99.2 | **93.2** | **98.6** | **99.3** | **83.9** | 99.5 | **77.3** | 98.7 | **95.8**<br>[95.3, 96.3] | **94.3**<br>[94.0, 94.6] |

Notably, despite mixed results in weakly supervised experiments, the ViT-L architecture seems to possess more inherent discriminative capacity. Indeed, we observe that combining ViT-L with the simplistic MeanPool MIL algorithm leads to an average improvement of 1.60% over the ViT-B architecture. This improvement rises to 2.40% on slide-level classification tasks and 3.3% on CRC-related tasks.

In a previous study conducted by (35), it is shown that large models may perform worse than smaller ones when a “small” pre-training dataset is used. Similarly, we observe a saturation phenomenon with the largest architecture, which could be attributed to overfitting on a relatively small organ-specific pre-trained dataset. However, the results using MeanPool on slide-level tasks or linear evaluation on patch-level tasks suggest that ViT-L possesses higher intrinsic discriminative capacity and produces features that are more linearly separable than for smaller architectures. This property, however, does not benefit when frozen features are combined with non-linear advanced MIL algorithms such as ABMIL or TransMIL.

Finally, it is worth noting that the ViT-L architecture performs worse overall when combined with the TransMIL algorithm (*−* 0.8% and *−* 1.2% compared to ViT-S and ViT-B architectures). This supports the observations from the previous section, suggesting that TransMIL’s performance decreases with the dimension of the output space.

### 5.3 Scaling iBOT with pan-cancer dataset

In the previous section, we highlighted performance saturation reached with a ViT-L architecture pre-trained on a relatively small pre-training dataset TCGA-COAD. Consequently, we now investigate whether increasing both the pre-training dataset size and diversity helps the discriminative feature learning of our iBOT models. Table 8 compares the same ViT-B pre-trained with iBOT on two different pre-training sets, TCGA-COAD and PanCancer40M (43.3M patches). On average across all tasks, pan-cancer pre-training brings a slight improvement of 0.5% with ABMIL (1.3% with TransMIL) over colon adenocarcinoma pre-training. Interestingly, pan-cancer pre-training appears beneficial even for classification tasks involving organs unseen during pre-training (0.6% and 1.6% with ABMIL and TransMIL, respectively). Notably, iBOT[ViT-B]PanCancer outperforms its TCGA-COAD counterpart on breast cancer HRD prediction (79.3 vs. 74.4 ROC AUC), MSI prediction in stomach cancer (89.9 vs. 84.3 ROC AUC) or OS prediction in pancreatic cancer (59.2 vs. 56.1 ROC AUC with TransMIL). Moreover, pan-cancer pre-training does not induce a major performance drop on colorectal cancer tasks compared to TCGA-COAD pre-training, except on PAIP-CRC external validation with a ROC AUC drop of 1.8 points (see Table 9). Thus, pan-cancer and colon adenocarcinoma pre-trainings act in a complementary fashion. These observations are also confirmed on patch classification tasks. iBOT[ViT-B]PanCancer consistently outperforms iBOT[ViT-B]COAD on breast cancer Camelyon17-WILDS dataset (see Table 10), showing remarkable label efficiency for less than 1% of the training dataset (see Figure 4). On the other hand, TCGA-COAD pre-training benefits patch-classification on NCT-CRC-HE-7K (as illustrated in Table 10) in all data regimes (Figure 4).

**Table 8.**
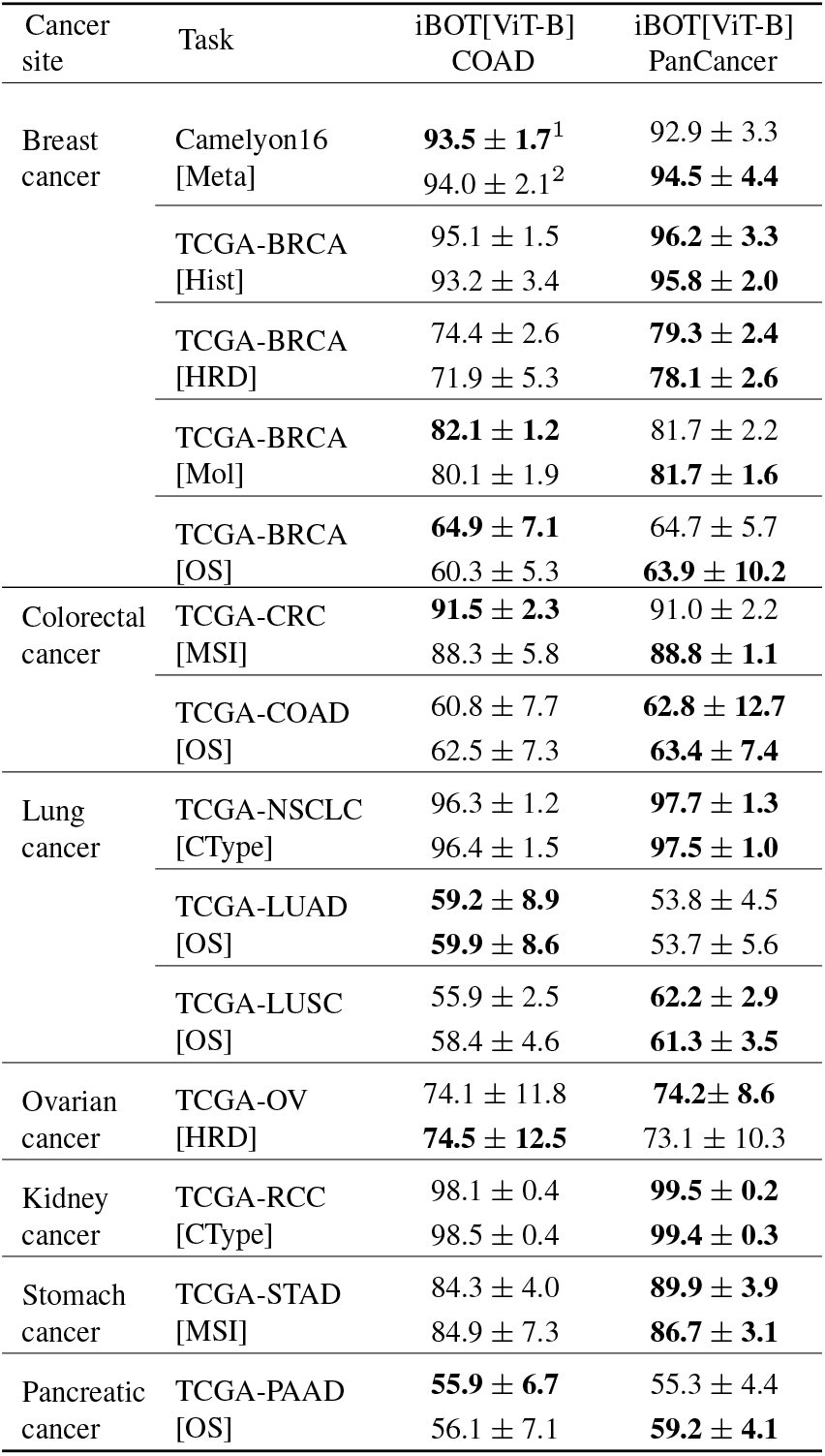
Impact of the pre-training dataset size on weakly-supervised downstream performance for a ViT-B architecture. The iBOT[ViT-B]COAD column is repeated from Table 2 to ease the comparison with iBOT[ViT-B]PanCancer. ROC AUC scores and C-Index are reported for classification and survival tasks, respectively. We take the average and standard deviation of each metric over the five outer test splits from nested CV. Bold indicates the highest performance for each MIL model (^1^ABMIL, ^2^ TransMIL).

**Table 9.**
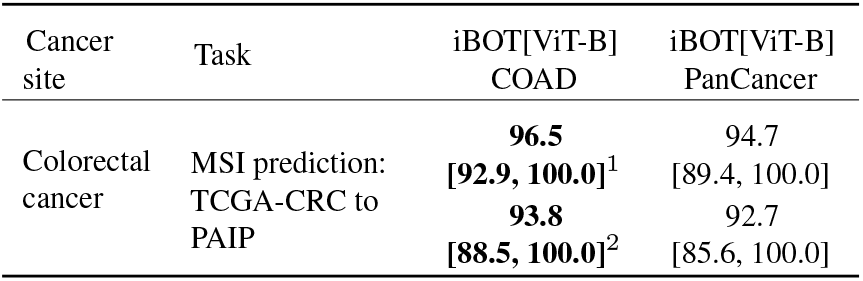
Performance comparison of iBOT ViT-B pre-trained on TCGA-COAD vs. PanCancer40M on PAIP-CRC[MSI] external validation. The iBOT[ViT-B]COAD column is repeated from Table 3 to ease the comparison with iBOT[ViT-B]PanCancer. ROC AUC scores and 95% confidence intervals are computed using bootstrap with 1,000 repeats. The top and bottom rows indicate performance with ABMIL^1^ and TransMIL^2^.

**Table 10.** Impact of ViT pre-training datasets on patch classification tasks performance for a ViT-B architecture. The iBOT[ViT-B]COAD line is repeated from Table 4. F1 score (*†*) is reported for single class classification (ADI to TUM) in NCT-CRC-HE-7K. Accuracy (*‡*) and 95% confidence intervals are computed using bootstrap with 1,000 repeats for multi-class classification in NCT-CRC-HE-7K and binary classification in Camelyon17-WILDS, respectively. Bold indicates the highest performance across classes. ROCAUC scores are reported in appendix (Table G3).

| Method | NCT-CRC-HE-7K |  |  |  |  |  |  |  |  | Camelyon<br>17WILDS |
| --- | --- | --- | --- | --- | --- | --- | --- | --- | --- | --- |
| | Adi† | Deb† | Lym† | Muc† | Mus† | Norm† | Str† | Tum† | All‡ | Metastases‡ |
| iBOT[ViT-B]<br>COAD | 99.4 | **92.7** | **96.6** | **99.0** | 81.1 | **99.6** | 75.8 | **98.9** | **95.3**<br>[94.8, 95.9] | 93.7<br>[93.4, 93.9] |
| iBOT[ViT-B]<br>PanCancer | **99.5** | 83.7 | 88.8 | **99.0** | **83.7** | 99.4 | **77.4** | 98.5 | 94.3<br>[93.7, 94.9] | **96.6**<br>[96.4, 96.8] |

**Fig. 4.**
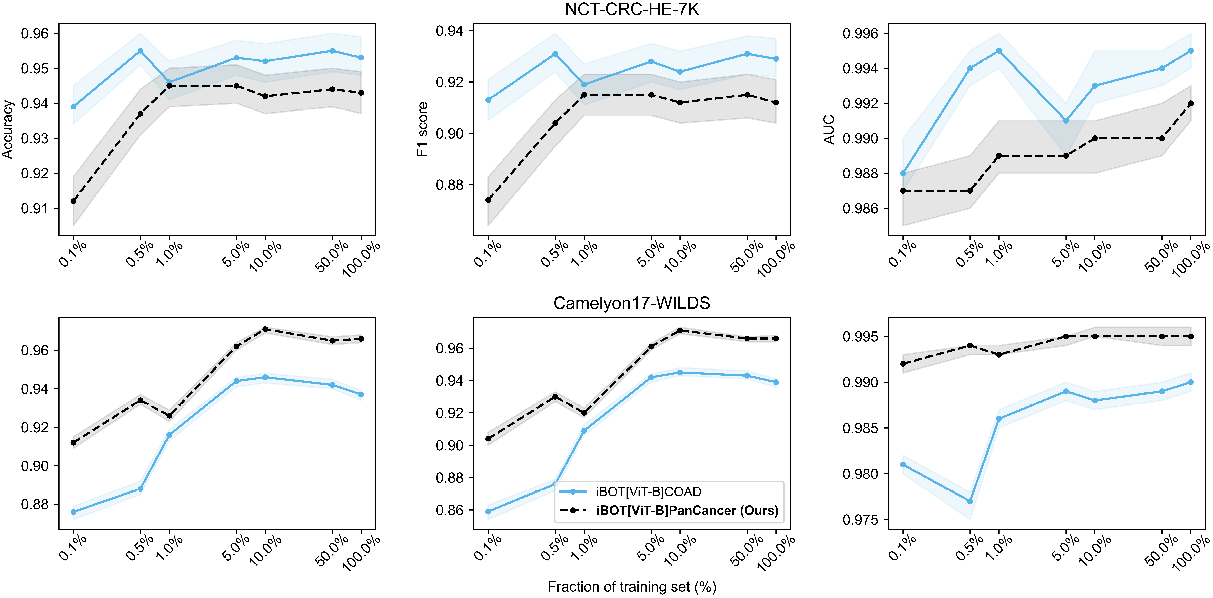
Impact of ViT pre-training datasets on the linear evaluation results with different sizes of training data for a ViT-B architecture. Results are reported on NCT-CRC-HE and Camelyon17-WILDS testing datasets. Metrics are reported for an ensemble of 30 linear classifiers with different initializations. 95% confidence intervals are computed using bootstrap with 1,000 repeats.

Overall and in contrast with (38), we conclude that pre-training on a larger, pan-cancer dataset often improves the performance in downstream tasks involving unseen cancer indications. Although no consistent performance drop on colon-specific tasks is observed for iBOT[ViT-B]PanCancer, colon-specific pre-training benefits better generalization, which should be further investigated with more internal and external validation cohorts.

According to the findings of (38), adding diversity to the pre-training dataset does not necessarily result in a more generalized model per se. As an additional study, we compare two ViT-S iBOT models that were pre-trained on TCGA-COAD and PanCancer4M datasets respectively, both following the same experimental setup and containing an equal number of tiles. We specifically investigate the impact of increasing the diversity of organ sites during pre-training. In appendix E, our results show an average performance drop of 1.3% from colon to pan-cancer pre-training, this across all weakly-supervised tasks using the ABMIL aggregation algorithm (2.4% with TransMIL, as shown in appendix, Table E2). Moreover, we notice a substantial decrease of 8.4 points in the ROC AUC score for the external validation of PAIP-CRC when considering a pan-cancer pre-training approach. This suggests that as diversity increases, the pre-training task becomes more challenging, particularly for the ViT-S model which has limited capacity to handle complex variations. However, it is worth noting that the iBOT[ViT-S]PanCancer model outperformed its TCGA-COAD counterpart in patch-classification tasks, demonstrating significant improvements across all data regimes (*p <* 0.005 for all datasets and training set ratios, except for the 5% ratio) (refer to Figure E1 in appendix). We speculate that a smaller model with restricted representation capacity benefits from higher diversity during pre-training from the perspective of learning more discriminative higher-level features. This attribute benefits simple tissue phenotyping tasks but faces limitations on more challenging tasks such as weakly-supervised classification.

In contrast, the iBOT[ViT-S]COAD iterating on a smaller set of patches, tends to focus more on specific histology characteristics during training. This narrower focus can simplify the training process and enhance the network’s representation capabilities. However, this approach also poses the risk of overfitting to colon-specific features and may limit the model’s ability to generalize and abstract information during linear evaluation tasks.

Together with the ViT-Base comparison, those results confirm the assumption that histology pre-training pipelines benefit from a simultaneous scaling of both the dataset size (in order to have more variety in terms of patches) and the network capacity (in order to increase its representation capabilities), as highlighted by Figure 5.

**Fig. 5.**
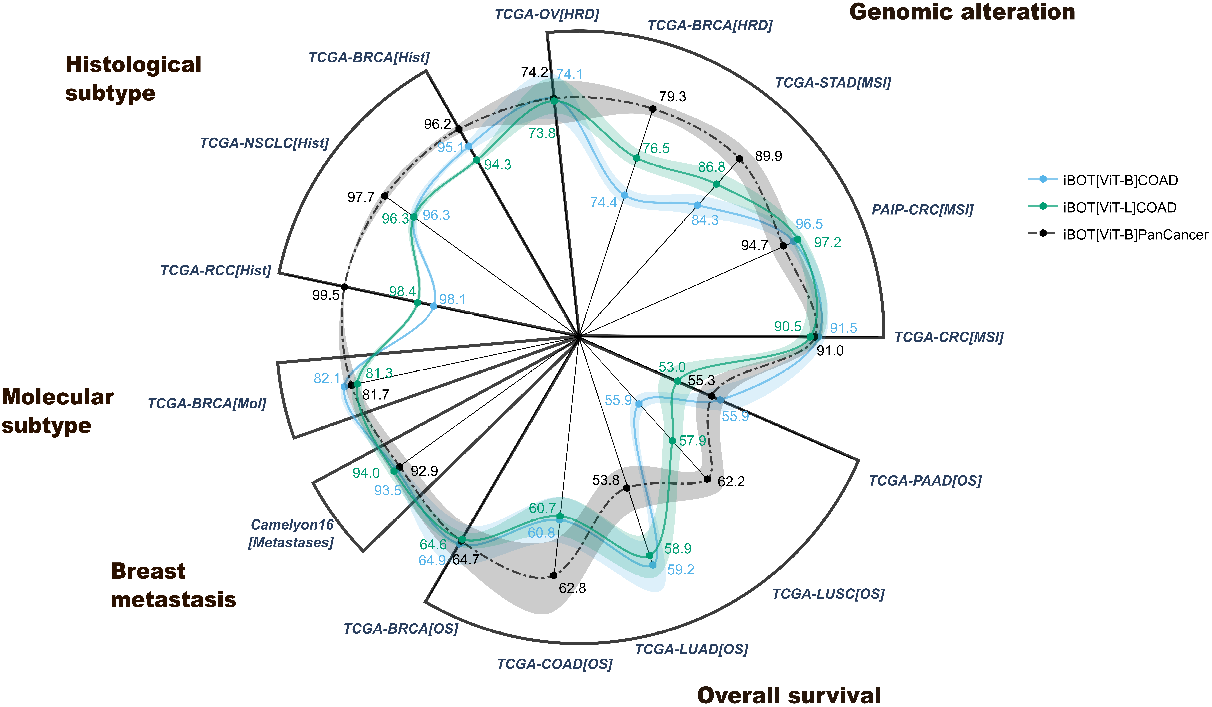
Scaling iBOT to 80M parameters with pan-cancer pre-training on more than 40M histology tiles. We report the performance obtained with ABMIL aggregation algorithm. The 5 × 5 nested CV is applied without repetition. We report the average test metrics and standard deviation on the outer folds. ROC AUC scores and Harrell’s C-Index ([OS] suffix) are shown for classification and survival tasks, respectively. CRC-specific tasks are highlighted in bold.

### 5.4 Comparison with other in-domain pre-trained methods

Eventually, this last section provides a comparison of iBOT[ViT-B]PanCancer with state-of-the-art SSL models used in computational pathology. Those include: i) a ViT pre-trained with knowledge distillation (Dino[ViT-S]BRCA and HIPT) or CL (MoCoV2[RN50W2]COAD); ii) hybrid CNN and transformer encoder framework pre-trained with semantically relevant CL, denoted by CTransPath (24).

Note that Figure 1 serves as a graphical summary of Table 11 and Table 12, which we detail hereinafter.

**Table 11.** Comparison of state-of-the-art SSL frameworks on weakly-supervised downstream performance. We display the best performance across five MIL algorithms for features aggregation: TransMIL, ABMIL, DSMIL, Chowder and MeanPool. A sixth transformer-based algorithm is considered for HIPT based on the original implementation. ROC AUC and C-Index are reported for classification and survival tasks, respectively. We take the average and standard deviation of each metric over the five outer test splits from nested CV. Bold and underline indicate the highest and second highest performance across SSL methods, respectively.

| Cancer site | Task | (A)<br>Cohort-specific pre-training |  | (B)<br>Pan-cancer pre-training |  |  |
| --- | --- | --- | --- | --- | --- | --- |
|  |  | Dino[ViT-S]<br>BRCA | MoCoV2<br>[RN50W2]<br>COAD | HIPT | CTransPath | iBOT[ViT-B]<br>PanCancer |
| Breast cancer | Camelyon16[Meta] | 84.5 ± 4.0 | 91.4 ± 4.3 | <u>95.7 ± 2.1</u> | **96.3 ± 2.6** | 94.5 ± 2.8 |
|  | TCGA-BRCA[Hist] | 92.1 ± 3.0 | 93.0 ± 1.7 | 91.3 ± 1.9 | <u>95.8 ± 0.5</u> | **96.2 ± 1.5** |
|  | TCGA-BRCA[HRD] | 72.1 ± 3.1 | 73.5 ± 4.3 | 73.1 ± 3.9 | <u>77.1 ± 2.5</u> | **79.3 ± 2.7** |
|  | TCGA-BRCA[Mol] | 77.9 ± 1.9 | 78.0 ± 1.4 | 78.9 ± 3.4 | <u>80.8 ± 1.7</u> | **81.7 ± 1.6** |
|  | TCGA-BRCA[OS] | 60.3 ± 10.2 | 62.6 ± 7.0 | 63.9 ± 5.8 | **65.0 ± 6.0** | <u>64.7 ± 5.7</u> |
| Colorectal cancer | TCGA-CRC[MSI] | 76.1 ± 4.4 | 88.5 ± 2.5 | 83.1 ± 4.3 | <u>88.5 ± 2.3</u> | **91.0 ± 2.2** |
|  | TCGA-COAD[OS] | 57.7 ± 10.4 | 62.6 ± 9.3 | 60.6 ± 3.4 | **64.3 ± 5.4** | <u>63.4 ± 7.4</u> |
| Lung cancer | TCGA-NSCLC[CType] | 92.8 ± 2.5 | 96.2 ± 1.7 | 94.2 ± 2.8 | <u>97.3 ± 0.4</u> | **97.7 ± 1.3** |
|  | TCGA-LUAD[OS] | 59.1 ± 4.1 | **61.6 ± 2.9** | 58.3 ± 3.0 | <u>59.1 ± 4.5</u> | 58.0 ± 6.8 |
|  | TCGA-LUSC[OS] | 60.8 ± 4.0 | <u>61.6 ± 4.2</u> | 61.1 ± 5.7 | 61.5 ± 2.9 | **63.2 ± 1.4** |
| Ovarian cancer | TCGA-OV[HRD] | 57.2 ± 8.2 | <u>70.2 ± 11.4</u> | 69.5 ± 12.9 | 69.5 ± 7.0 | **74.2 ± 8.6** |
| Kidney cancer | TCGA-RCC[CType] | 97.5 ± 0.8 | 98.6 ± 0.3 | 98.6 ± 0.4 | <u>98.9 ± 0.2</u> | **99.5 ± 0.2** |
| Stomach cancer | TCGA-STAD[MSI] | 76.5 ± 3.3 | 79.0 ± 4.0 | 79.6 ± 3.1 | <u>83.2 ± 8.1</u> | **89.9 ± 3.9** |
| Pancreatic cancer | TCGA-PAAD[OS] | 59.3 ± 6.8 | <u>59.6 ± 4.2</u> | **61.3 ± 2.7** | 59.0 ± 4.2 | 59.2 ± 4.1 |

**Table 12.**
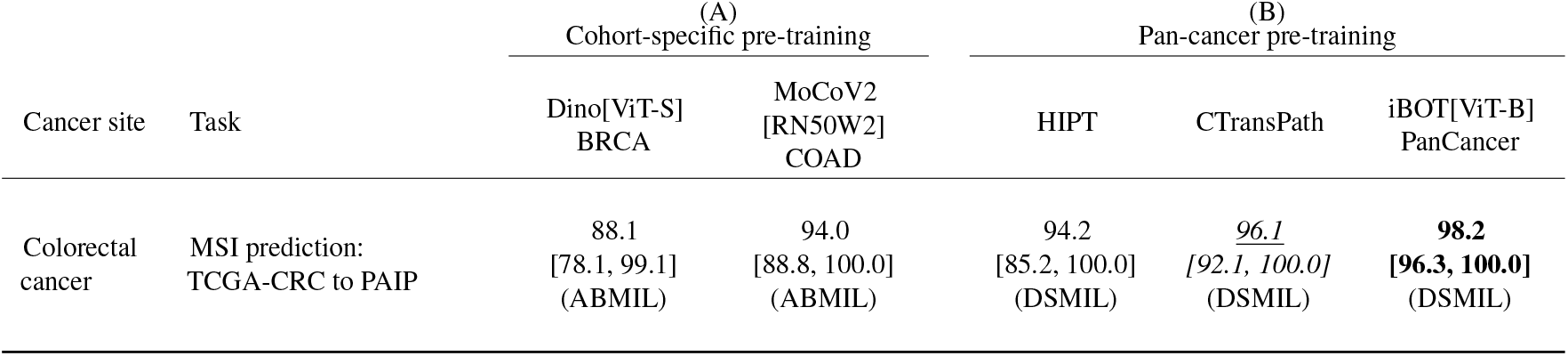
Comparison of state-of-the-art SSL frameworks on PAIP-CRC[MSI] external validation. Best MIL model is reported in parentheses. Bold and underline indicate the highest and second highest performance. ROC AUC scores and 95% confidence intervals are computed using bootstrap with 1,000 repeats.

| Cancer site | Task | (A)<br>Cohort-specific pre-training |  | (B)<br>Pan-cancer pre-training |  |  |
| --- | --- | --- | --- | --- | --- | --- |
|  |  | Dino[ViT-S]<br>BRCA | MoCoV2<br>[RN50W2]<br>COAD | HIPT | CTransPath | iBOT[ViT-B]<br>PanCancer |
| Colorectal cancer | MSI prediction:<br>TCGA-CRC to PAIP | 88.1<br>[78.1, 99.1]<br>(ABMIL) | 94.0<br>[88.8, 100.0]<br>(ABMIL) | 94.2<br>[85.2, 100.0]<br>(DSMIL) | <u>96.1</u><br>[92.1, 100.0]<br>(DSMIL) | **98.2**<br>**[96.3, 100.0]**<br>(DSMIL) |

**Table 13.**
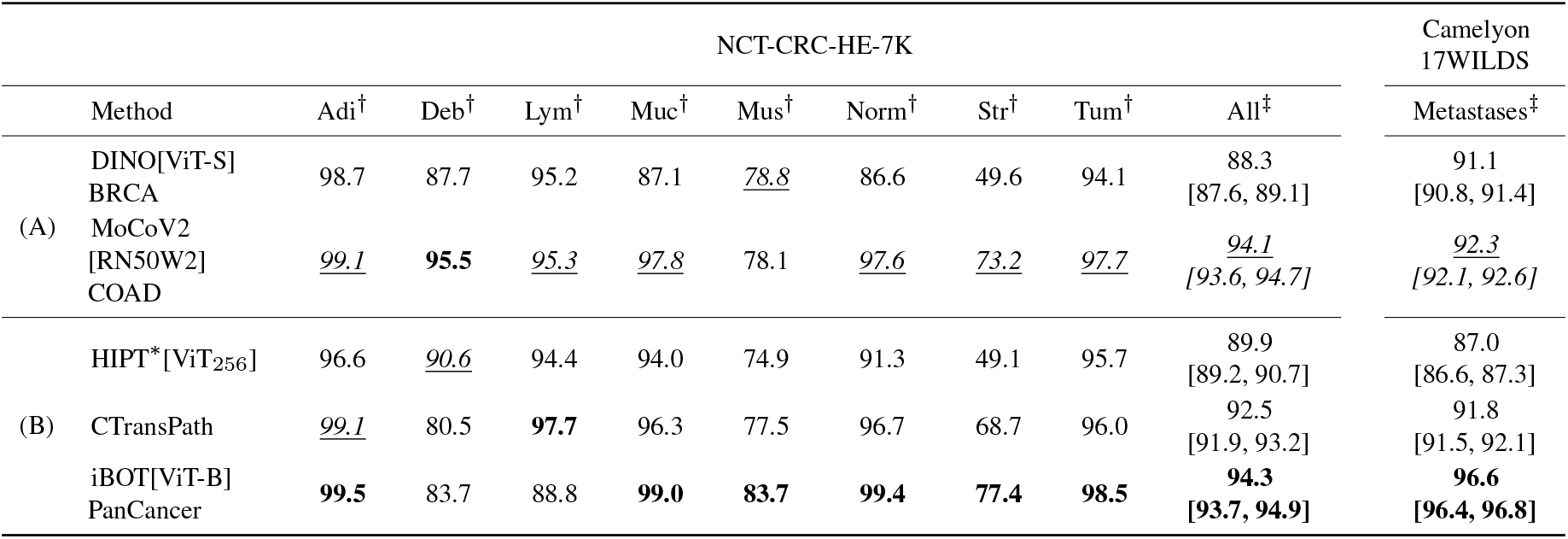
Comparison of state-of-the-art SSL frameworks on patch classification tasks. HIPT^*^[ViT256] correspond to the first ViT-S model of HIPT architecture pre-trained on 256 × 256 pixels tiles. F1 score (†) is reported for single class classification (Adi to Tum) in NCT-CRC-HE-7K. Accuracy (‡) and 95% confidence intervals are computed using bootstrap with 1,000 repeats for multi-class classification in NCT-CRC-HE-7K and binary classification in Camelyon17-WILDS, respectively. Bold and underline indicate the highest and second highest performance across classes, respectively. ROC AUC scores are reported in appendix (Table G4).

Table 11 displays, for each model, the maximal performance obtained across five different MIL models: TransMIL, ABMIL, DSMIL, Chowder and MeanPool. A sixth transformer-based algorithm is considered for HIPT based on the original implementation. We show that iBOT[ViT-B]PanCancer outperforms most other methods on 9 over 14 tasks with ABMIL aggregation model (see Table F1 in appendix for results with ABMIL). Our model pre-trained on pan-cancer data brings a 1.4% and 4.0% mean improvement on CTransPath and HIPT, respectively.

Among those three pan-cancer feature extractors, iBOT[ViT-B]PanCancer places first and achieves an average 1.2% (6.5%) gain on CRC-related tasks compared to CTransPath (resp. HIPT), and an average 1.5% (3.4%) gain on other tasks. Our model shows state-of-the-art performance on TCGA-RCC and TCGA-NSCLC histological subtype classification tasks, along with a remarkable generalization ROC AUC score on PAIP-CRC MSI prediction (see Table 12). In addition, iBOT[ViT-B]PanCancer depicts the higher performance on patch-classification tasks with full-training sets 13 surpassing other models by a large margin on Camelyon17-WILDS dataset. Low data regime scenarios depict on-par generalization performance between our model, CTransPath and MoCoV2[RN50W2]COAD, NCT-CRC-HE-7K dataset, with very high label efficiency on 0.1% of Camelyon17-WILDS training set (Figure 6). The previous results demonstrate the validity and superiority of our method in capturing high-level semantic features for patch phenotyping tasks, while producing highly informative features for intricate slide-level predictions.

**Fig. 6.**
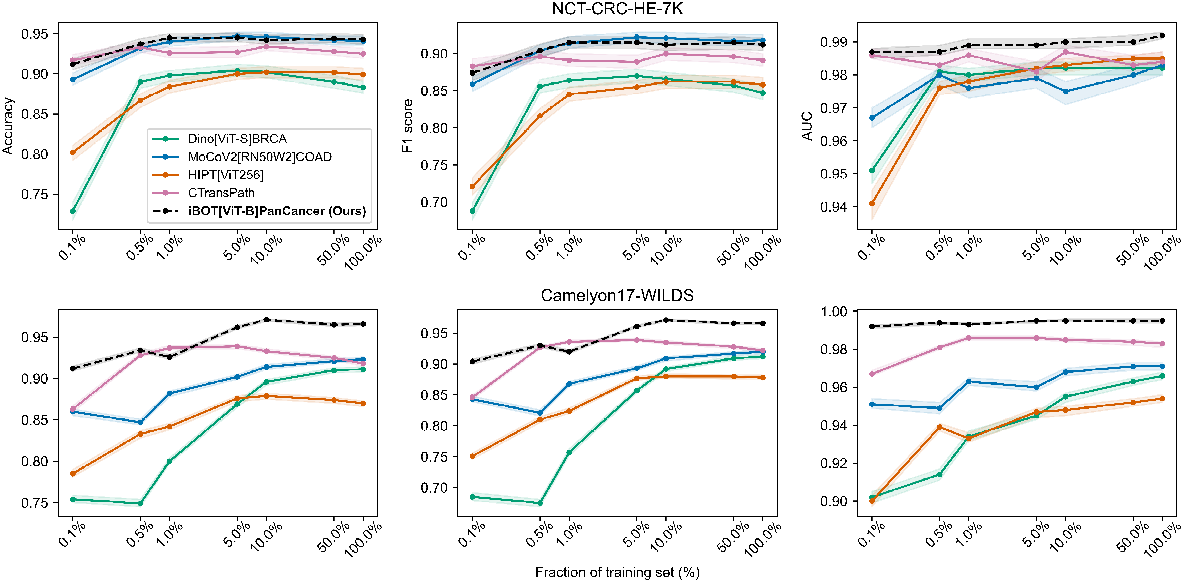
Linear evaluation results on NCT-CRC-HE and Camelyon17-WILDS testing dataset for different state-of-the-art SSL frameworks with increasing size of training data. Metrics are reported for an ensemble of 30 linear classifiers with different initializations. 95% confidence intervals are computed using bootstrap with 1,000 repeats.

## 6 Discussion

In this work, we explored the benefits of using iBOT to pre-train large neural networks on databases of unlabeled WSI. Through a large panel of 17 downstream tasks spanning seven cancer indications and 16 cancer subtypes, we showed that our iBOT ViT-B model, pre-trained on more than 40M histology patches, strongly improves the performance on ten weakly-supervised WSI classification compared to other SSL frameworks available in the literature. These results are based on an analysis of the learning scalability of iBOT both in terms of pre-training dataset size, pre-training dataset diversity and model architecture. As a result, we provide a set of guidelines for pre-training large ViT models on histology data using iBOT:

1. In-domain pre-training of ViT with iBOT should be favored over standard CNN pre-training on ImageNet. We show that a ViT pre-trained with iBOT outperforms a ResNet pre-trained on ImageNet on 16 out of 17 downstream tasks.
2. ViT pre-trained with iBOT should be favored over a standard CNN pre-trained with MoCo v2. We show that, when both pre-trained on TCGA-COAD, a ViT-B (86M parameters) pre-trained with iBOT outperforms a ResNet50-w2 pre-trained with MoCo v2 (67M) pre-trained with MoCo v2. Besides, a ViT-S model pre-trained with iBOT achieves on-par downstream performance when compared to the aforementioned ResNet model, with three times fewer parameters.
3. Pre-training on a relatively small histology dataset (4M patches) and increasing the size of the architecture to a ViT-B (85M parameters) yields consistent performance improvement over a ViT-S (21M parameters). However, increasing the size of the architecture to a ViT-L (307M parameters) does not yield further performance improvement.
4. In-cancer-domain pre-training benefits from a simultaneous scaling of both the dataset size, diversity and network capacity, with improved downstream generalization performance. Notably, increasing the sole diversity of the pre-training dataset from colon-specific to pan-cancer (PanCancer4M) seems to degrade the downstream performance of a ViT-S model pre-trained with iBOT. In contrast, a ViT-B model (86M parameters) pre-trained with 40M pan-cancer histology tiles outperform both a ViT-B and ViT-L (307M) models pre-trained with 4M colon-specific histology tiles on a wide variety of weakly-supervised tasks, without performance drop on colorectal cancer tasks.

In view of our experiments, we note that the above-mentioned guidelines need further validation. As such, a comprehensive ablation study should be conducted to disentangle the role of model architecture, number of pre-training iterations and pre-training dataset size with iBOT for histology images (35). Those experiments non-exclusively involve: (i) pre-training a ViT-L on the PanCancer40M dataset (which we estimate to take 11,000 V100 hours for 350,000 iterations), confirming that larger pan-cancer pre-training benefits larger models; (ii) pre-training a ViT-S PanCancer on 40M tiles, confirming that larger pan-cancer datasets are not required for smaller architectures like ViT-S; (iii) implementing a validation loss or custom metric (79), as it has been demonstrated to have a strong correlation with downstream performance (35), thus avoiding the need for expensive evaluation on weakly-supervised tasks. Lastly, our downstream evaluation protocol could be even further extended by broadening the list of downstream tasks, including segmentation, object detection, and retrieval tasks.

In section 4.1, we highlighted the superiority of a ViT-B model pre-trained with iBOT over a ResNet50-w2 trained with MoCo V2. Additional experiments should enrich this comparison, especially to decouple the SSL method (iBOT vs. MoCo v2/v3) and the pre-trained model (*i*.*e*., ViT vs. CNN). Moreover, the cross-entropy loss on <monospace>[CLS] </monospace>tokens was shown to be responsible for acquiring most of the visual semantics as a form of CL without positive pairs (33). This leaves room for further enhancing the CL component of iBOT. As such, one could enforce iBOT to further spread apart features in the output space, *e*.*g*., by replacing the standard cross-entropy loss on <monospace>[CLS] </monospace>tokens by InfoNCE loss (80) or KoLeo regularization (81, 82). In addition, the construction of positive and negative pairs tailored for histology could also be investigated, such as enforcing spatial proximity of positive pairs (MoCo v3).

It should also be noted that the downstream performance of SSL frameworks on weakly-supervised tasks remains dependent on the application and data at hand. In particular, OS prediction exhibits the highest variability across datasets with no clear trend between SSL frameworks. The OS label is known to show a limited correlation with histology features. It is important to note that patients with cancer may experience mortality from causes unrelated to cancer itself. This factor could contribute to the increased variability observed in model benchmarks for OS prediction tasks.

Even though our ViT-B model pre-trained iBOT on 43M pan-cancer patches demonstrates remarkable improvement over other SSL methods, pre-training ViT-models with iBOT is highly computationally intensive and may exhibit instability, especially for larger architectures and datasets. To address this issue, we intend to explore engineering enhancements to stabilize and speed up the pre-training process, making it more scalable to increasing model and dataset sizes. A recent study in computer vision suggests that such technical improvements have the potential to achieve a two-fold increase in speed and a three-fold gain in memory efficiency compared to the iBOT method (81). In combination with the validation loss mentioned above, these gains pave the way for further hyperparameters exploration. Lastly, our iBOT methods are pre-trained on histology tiles with the same data augmentation as the one used for natural images (ImageNet). Consequently, we could expect better generalization performance by integrating histology-specific data augmentation and normalization methods into the MIM framework (24, 83, 84).

## 7 Conclusion

In this work, we successfully scaled iBOT pre-training with large ViT models to massive datasets of unlabeled WSI. Our findings indicated that further scaling beyond ViT-B architectures offers the potential for the development of a foundation model for digital pathology. However, it is essential to acknowledge that scaling per se should not overlook the crucial role of data curation for SSL pre-training. We strongly believe that significant advancements can also be achieved by constructing a highly curated and balanced dataset that extends beyond TCGA WSI. Exciting avenues for improvement include the adoption of more efficient data sampling strategies (81) or the incorporation of automatic data subset selection during pre-training (85).

## Data Availability

The results published in this work are partly based upon data generated by the TCGA Research Network (TCGA). All images and the associated clinical outcome for TCGA cohorts used in this study are publicly available at https://portal.gdc.cancer.gov/ and cBioPortal https://www.cbioportal.org/. Regarding the PAIP dataset, de-identified pathology images and annotations used in this research were prepared and provided by the Seoul National University Hospital by a grant of the Korea Health Technology R&D Project through the Korea Health Industry Development Institute (KHIDI), funded by the Ministry of Health & Welfare, Republic of Korea (grant number: HI18C0316).

https://portal.gdc.cancer.gov/

https://www.cbioportal.org/

http://www.wisepaip.org/paip

## Credit authorship contribution statement

**Alexandre Filiot**Conceptualization, Formal analysis, Methodology, Software, Validation, Visualization, Writing - Original Draft, Reviewing & Editing. **Ridouane Ghermi**: Conceptualization, Investigation, Methodology, Software, Writing - Original Draft **Antoine Olivier**: Investigation, Methodology, Software, Writing - Reviewing & Editing. **Paul Jacob**: Investigation, Methodology, Software, Writing - Reviewing & Editing. **Lucas Fidon**: Investigation, Methodology, Software, Writing - Reviewing Editing. **Alice Mac Kain**: Funding acquisition, Project administration, Writing - Reviewing & Editing. **Charlie Saillard**: Conceptualization, Investigation, Methodology, Software, Supervision, Writing - Reviewing & Editing. **Jean-Baptiste Schiratti**: Conceptualization, Methodology, Software, Supervision, Writing - Reviewing & Editing.

## Acknowledgements

This work was granted access to the High-Performance Computing (HPC) resources of IDRIS under the allocations 2022-AD011012519 and 2023-AD011012519R1 made by GENCI.

The results published here are partly based upon data generated by the TCGA Research Network: https://www.cancer.gov/tcga. Regarding the PAIP dataset, de-identified pathology images and annotations used in this research were prepared and provided by the Seoul National University Hospital by a grant of the Korea Health Technology R&D Project through the Korea Health Industry Development Institute (KHIDI), funded by the Ministry of Health & Welfare, Republic of Korea (grant number: HI18C0316).

We thank Benjamin Adjadj and Auriane Riou for their valuable contribution to the early development of our methodology, as well as the investigation of additional experiments. We thank Jean-Philippe Vert and Eric Durand for their detailed proofreading and insightful comments.

## Supplementary Material

Supplementary material are given as a separate document.

## Footnotes

1 In this document, we use the “4M” notation to denote “4 million”.

2 See https://github.com/owkin/HistoSSLscaling.

