## Supplementary Material for "Scaling Self-Supervised Learning for Histopathology with Masked Image Modeling"

### Appendix A: iBOT pre-training parameters

In addition to hyperparameters described in section 3.3, iBOT models were pre-trained using weight decay, a shared projection head with output dimension 8,192 and gradient clipping. We provide details on each of these hyperparameters.

- **Projection head**: for all Vision Transformers (ViT), the projection head consists of a 3-layer multi-layer perceptron (MLP) with hidden dimension 2,048, followed by L2 normalization and a weight normalized fully connected layer with output dimension 8,192. Based on the empirical results presented in (1), the projection head was shared between the [CLS] token and patch tokens, with output dimension 8,192. Finally, the last linear layer of the projection head was kept frozen for the first three epochs to ensure convergence.
- **Weight decay**: initial value of weight decay was set to 0.04, with a final value of 0.4 for ViT-S/16 and ViT-B/16, 0.48 for ViT-L/16. A cosine schedule is applied.
- **Gradient clipping**: gradient clipping was applied with a value of 3 (*i.e.* all gradients have a maximal L2 norm of 3) to all ViT models.

### Appendix B: Repartition of pan-cancer pre-training datasets

Two pan-cancer pre-training datasets are used in this study: PanCancer40M and PanCancer4M. PanCancer40M pre-training dataset covers 13 anatomic sites and 16 cancer subtypes for 5,558 patients, representing a total of 6,093 slides and 43,374,634 patches. PanCancer4M is taken as a subset (random sampling) of its larger counterpart, with 5,183 whole slide images (WSI) and 4,386,755 tiles overall. We provide additional details on the distribution of The Cancer Genome Atlas (TCGA) cohorts for each of these two datasets (see Table B1 and Table B2 below).

**Table B1.** Distribution of TCGA cohorts in PanCancer40M pre-training dataset.

| Cohort | No. patients | No. WSIs | No. patches |
|---|---|---|---|
| TCGA-KIRC | 514 | 519 | 1,299,576 |
| TCGA-KICH | 121 | 121 | 1,299,903 |
| TCGA-PAAD | 189 | 209 | 3,197,302 |
| TCGA-COAD | 434 | 441 | 3,499,776 |
| TCGA-READ | 157 | 158 | 499,912 |
| TCGA-OV | 107 | 107 | 1,950,937 |
| TCGA-LUSC | 478 | 512 | 1,999,872 |
| TCGA-LUAD | 468 | 530 | 1,999,531 |
| TCGA-PRAD | 278 | 310 | 3,984,428 |
| TCGA-BLCA | 386 | 457 | 3,999,664 |
| TCGA-BRCA | 1,060 | 1,124 | 3,999,192 |
| TCGA-UCEC | 506 | 566 | 3,999,922 |
| TCGA-KIRP | 107 | 110 | 1,293,676 |
| TCGA-LIHC | 363 | 371 | 3,999,751 |
| TCGA-ESCA | 15 | 158 | 2,358,538 |
| TCGA-STAD | 375 | 400 | 3,992,654 |
| All | 5,558 | 6,093 | 43,374,634 |

**Table B2.** Distribution of TCGA cohorts in PanCancer4M pre-training dataset.

| Cohort | No. patients | No. WSIs | No. patches |
|---|---|---|---|
| TCGA-KIRC | 514 | 519 | 129,750 |
| TCGA-KICH | 121 | 121 | 129,954 |
| TCGA-PAAD | 189 | 209 | 399,817 |
| TCGA-COAD | 434 | 441 | 349,713 |
| TCGA-READ | 157 | 158 | 49,928 |
| TCGA-OV | 107 | 107 | 399,966 |
| TCGA-LUSC | 478 | 512 | 199,680 |
| TCGA-LUAD | 468 | 530 | 199,810 |
| TCGA-PRAD | 278 | 310 | 399,900 |
| TCGA-BLCA | 386 | 457 | 399,875 |
| TCGA-BRCA | 1,060 | 1,124 | 399,020 |
| TCGA-UCEC | 506 | 566 | 399,596 |
| TCGA-KIRP | 107 | 110 | 129,910 |
| TCGA-LIHC | 363 | 371 | 399,938 |
| TCGA-ESCA | 15 | 158 | 399,898 |
| All | 5,183 | 5,693 | 4,386,755 |

### Appendix C: SSL methods

Table C1 provides a descriptive summary of each of the representation learning frameworks used as feature extraction methods in this study. Following section 4.3, models are named using the framework-architecture-dataset formalism. The weights from Dino[ViT-S]BRCA (2), HIPT (3) and CTransPath (4) models were retrieved directly from their respective GitHub repositories.

**Table C1.** Description of the representation learning frameworks considered in this study. Learning paradigms: Supervised Learning (SL) and Self-Supervised Learning (SSL). Domains: Out-Of-Domain (OOD), such as ImageNet-1K, and In-Domain (ID), such as histology datasets. SSL paradigms: contrastive learning (CL), Semantically-Relevant Contrastive Learning (SRCL), Knowledge Distillation (KD) and Masked Image Modeling (MIM).

| Model name | Learning paradigm | Domain | SSL paradigm | Model | No. params | Size of pre-trained datasets |
|---|---|---|---|---|---|---|
| Sup[RN50]IN | SL | OOD | - | RN50 | 23.5M | 1.2M |
| MoCoV2[RN50W2]COAD | SSL | ID | CL | RN50w2 | 66.8M | 4.4M |
| CTransPath | SSL | ID | (SR)CL | Swin-T + CNN | 27.5M | 14.3M |
| HIPT | SSL | ID | KD | ViT-S/16 | 21.7M | 104M |
| | | | | ViT-XS/256 | 2.8M | 0.4M |
| Dino[ViT-S]BRCA | SSL | ID | KD | ViT-S/16 | 21.7M | 2.1M |
| iBOT[ViT-S]COAD | SSL | ID | MIM | ViT-S/16 | 21.7M | 4.4M |
| iBOT[ViT-B]COAD | SSL | ID | MIM | ViT-B/16 | 85.8M | 4.4M |
| IBOT[ViT-L]COAD | SSL | ID | MIM | ViT-L/16 | 307M | 4.4M |
| iBOT[ViT-S]PANCAN | SSL | ID | MIM | ViT-S/16 | 21.7M | 43.3M |
| iBOT[ViT-B]PANCAN | SSL | ID | MIM | ViT-B/16 | 86M | 43.3M |

### Appendix D: Weakly-supervised tasks on TCGA cohorts

Table D1 provides an extensive description of TCGA cohorts and corresponding downstream tasks used throughout this study.

**Table D1.** TCGA cohorts and corresponding weakly-supervised slide-level downstream tasks.

| TCGA cohort | Cancer type | Task | Classes | No. patients | No. slides | Distribution |
|---|---|---|---|---|---|---|
| BRCA | Breast invasive carcinoma | Histological subtype classification | IDC<br>ILC | 938 | 1.001 | 79.8% (IDC)<br>20.2% (ILC) |
| | | Molecular subtype classification | Basal<br>Her2<br>LumA<br>LumB<br>Normal | 939 | 1,005 | 17.2% (Basal)<br>7.6% (Her2)<br>49.9% (LumA)<br>21.9% (LumB)<br>3.4% (Normal) |
| | | HRD prediction | HRD-L<br>HRD-H | 1,003 | 1,073 | 80.4% (HRD-L)<br>19.6% (HRD-H) |
| | | OS prediction | Continuous | 105 | 1,122 | Censoring: 86.7% |
| COAD | Colon adenocarcinoma | OS prediction | Continuous | 431 | 450 | Censoring: 78.0% |
| CRC | Colorectal carcinoma | MSI prediction | MSS/MSI-L<br>MSI-H | 555 | 576 | 85.6% (MSS/MSI-L)<br>14.4% (MSI-H) |
| LUAD | Lung adenocarcinoma | OS prediction | Continuous | 459 | 528 | Censoring: 60.2% |
| NSCLC | Non-small cell lung carcinoma | Cancer type classification | LUAD<br>LUSC | 947 | 1,05 | 51.1% (LUAD)<br>48.9% (LUSC) |
| OV | Ovarian serous cystadenocarcinoma | HRD prediction | HRD-L<br>HRD-H | 96 | 96 | Censoring: 50% |
| PAAD | Pancreatic adenocarcinoma | OS prediction | Continuous | 183 | 209 | Censoring: 45.9% |
| RCC | Kidney renal cell carcinoma | Histological subtype classification | KIRC<br>KIRP<br>KICH | 882 | 934 | 56.6% (KIRC)<br>30.4% (KIRP)<br>13.0% (KICH) |
| STAD | Stomach adenocarcinoma | MSI prediction | MSS/MSI-L<br>MSI-H | 375 | 401 | 83.9% (MSS/MSI-L)<br>16.2% (MSI-H) |

### Appendix E: Increasing dataset diversity for iBOT ViT-S pre-training

As an additional study, we compare two ViT-S models pre-trained with iBOT both on the colon adenocarcinoma cohort of TCGA (TCGA-COAD) and PanCancer4M datasets. We specifically investigate the impact of increasing the diversity of cancer indications during pre-training for a same number of tiles. This section encompasses all the results obtained to draw a fair comparison between our two models. Results analysis can be found in section 5.4 in the manuscript.

**Table E1-A.** Performance comparison of iBOT ViT-S pre-trained on TCGA-COAD vs. PanCancer4M for PAIP-CRC[MSI] external validation after training on TCGA-CRC[MSI] classification task. area under the receiver operating characteristic curve (ROC AUC) scores and $95\%$ confidence intervals are computed using bootstrap with $1,000$ repeats. Top and bottom row indicate performance with ABMIL[1] and TransMIL[2].

| Cancer site | Task | iBOT[ViT-S]COAD | iBOT[ViT-S]PanCancer |
|---|---|---|---|
| Breast cancer | Camelyon16 [Metastases] | 93.0 ± 5.8[1]<br>93.4 ± 4.7[2] | **93.4** ± **2.8**<br>**94.0** ± **3.9** |
| | TCGA-BRCA [Hist] | 94.0 ± 1.0<br>92.9 ± 2.0 | **95.2** ± **1.4**<br>**94.3** ± **1.6** |
| | TCGA-BRCA[HRD] | 72.8 ± 3.6<br>74.0 ± 3.5 | **74.7** ± **2.2**<br>**75.8** ± **2.9** |
| | TCGA-BRCA[Mol] | 79.4 ± 1.3<br>79.9 ± 2.0 | **80.0** ± **3.2**<br>**81.6** ± **0.9** |
| | TCGA-BRCA[OS] | **62.9** ± **8.5**<br>**63.8** ± **8.4** | 61.7 ± 6.4<br>62.2 ± 4.0 |
| Colorectal cancer | TCGA-CRC [MSI] | **89.1** ± **3.1**<br>**88.3** ± **5.6** | 87.6 ± 4.2<br>88.8 ± 1.1 |
| | TCGA-COAD [OS] | 58.5 ± 9.8<br>**62.9** ± **8.0** | **59.5** ± **6.5**<br>56.3 ± 2.7 |
| Lung cancer | TCGA-NSCLC [CancerType] | 94.7 ± 1.9<br>94.9 ± 3.0 | **97.2** ± **1.2**<br>**96.5** ± **1.5** |
| | TCGA-LUAD [OS] | **58.4** ± **5.2**<br>**59.3** ± **7.4** | 57.0 ± 5.9<br>57.5 ± 7.3 |
| | TCGA-LUSC [OS] | 57.7 ± 2.1<br>57.2 ± 5.9 | **58.2** ± **3.2**<br>**61.4** ± **2.6** |
| Ovarian cancer | TCGA-OV [HRD] | **72.2** ± **12.6**<br>**71.0** ± **4.8** | 65.8 ± 7.7<br>60.4 ± 10.1 |
| Kidney cancer | TCGA-RCC [CancerType] | 98.5 ± 0.5<br>98.3 ± 0.6 | **99.0** ± **0.3**<br>**98.8** ± **0.5** |
| Stomach cancer | TCGA-STAD [MSI] | **79.5** ± **3.8**<br>**82.5** ± **4.2** | 76.7 ± 5.4<br>80.6 ± 5.9 |
| Pancreatic cancer | TCGA-PAAD [OS] | **55.2** ± **3.6**<br>**57.7** ± **4.9** | 51.8 ± 7.8<br>53.0 ± 5.7 |

**Table E1-B.** Performance comparison of iBOT ViT-S pre-trained on TCGA-COAD vs. PanCancer4M for PAIP-CRC[MSI] external validation after training on TCGA-CRC[MSI] classification task. ROC AUC scores and $95\%$ confidence intervals are computed using bootstrap with $1,000$ repeats. Top and bottom row indicate performance with ABMIL[1] and TransMIL[2].

| Cancer site | Task | iBOT[ViT-S]COAD | iBOT[ViT-S]PanCancer |
|---|---|---|---|
| Colorectal cancer | MSI prediction<br>TCGA-CRC to PAIP | **96.5**<br>**[92.9, 100.0]**[1]<br><br>94.7<br>[89.4, 100.0] | **93.8**<br>**[88.5, 100.0]**[2]<br><br>92.7<br>[85.6, 100.0] |

**Table E2.** Impact of ViT pre-training dataset on patch classification tasks performance. F1 score (†) is reported for single class classification (Adi to Tum) in NCT-CRC-HE-7K. Accuracy (‡) and $95\%$ confidence intervals are computed using bootstrap with $1{,}000$ repeats for multi-class classification in NCT-CRC-HE-7K and binary classification in Camelyon17-WILDS, respectively. Bold indicates the highest performance across classes.

| | NCT-CRC-HE-7K | | | | | | | | | Camelyon17-WILDS |
|---|---|---|---|---|---|---|---|---|---|---|
| Method | Adi† | Deb† | Lym† | Muc† | Mus† | Norm† | Str† | Tum† | All‡ | Metastases‡ |
| iBOT[ViT-S]COAD | 99.5 | 97.6 | 98.0 | 99.6 | 96.4 | **99.7** | **96.1** | **99.4** | 93.2 [92.6, 93.8] | 92.4 [92.2, 92.7] |
| iBOT[ViT-S]PanCancer | **99.8** | **99.0** | **99.7** | **99.8** | **96.3** | **99.7** | 96.0 | 99.3 | **94.8** **[94.2, 95.3]** | **93.8** **[93.6, 94.0]** |

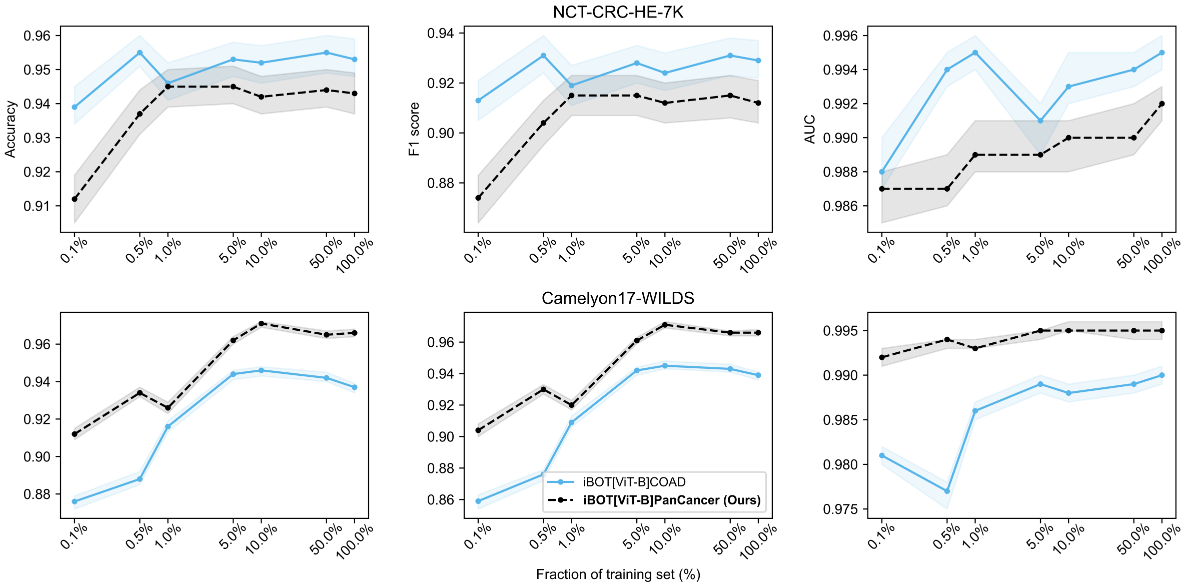


**Fig. E3.** Impact of ViT pre-training datasets on the linear evaluation results with different sizes of training data for a ViT-S architecture. Results are reported on NCT-CRC-HE and Camelyon17-WILDS testing dataset. Metrics are reported for an ensemble of $30$ linear classifiers with different initializations. $95\%$ confidence intervals are computed using bootstrap with $1{,}000$ repeats.

### Appendix F: Comparison with other in-domain SSL methods

In addition to section 5.4, we provide the downstream performance of Dino[ ViT-S]BRCA, MoCoV2[RN50W2]COAD, HIPT, CTransPath and iBOT[ViT-B]PanCancer on slide-level tasks using ABMIL aggregation method specifically.

**Table F1.** Comparison of state-of-the-art SSL frameworks on weakly-supervised downstream tasks. We display the performance with ABMIL. Results are reported for a set of 14 weakly-supervised prediction tasks across seven cancer indications. Bold indicates the highest performance across SSL models and multiple instance learning (MIL) models. [MSI], [HRD], [Ctype], [Mol], [Hist] and [OS] denote respectively: Microsatellite Instability (MSI), Homologous Recombination Deficiency (HRD), Cancer Type, Molecular Subtyping, Histological Subtyping classification, and overall survival (OS) prediction. ROC AUC scores and C-Index are reported for classification and survival tasks, respectively. We take the average and standard deviation of each metric over the five outer test splits from nested cross-validation (CV).

| | | (A) Cohort-specific pre-training | | (B) Pan-cancer pre-training | | |
|---|---|---|---|---|---|---|
| Cancer site | Task | Dino[ViT-S] BRCA | MoCoV2 [RN50W2] COAD | HIPT | CTrans Path | iBOT[ViT-B] PanCancer |
| Breast cancer | Camelyon16[Meta] | 83.8 ± 4.3 | 91.4 ± 4.3 | **95.7 ± 1.3** | 93.9 ± 4.4 | 92.9 ± 3.3 |
| | TCGA-BRCA[Hist] | 92.1 ± 3.0 | 93.0 ± 1.7 | 91.3 ± 1.9 | 95.4 ± 0.8 | **96.2 ± 1.5** |
| | TCGA-BRCA[HRD] | 72.1 ± 3.1 | 73.5 ± 4.3 | 73.1 ± 3.5 | 76.8 ± 2.9 | **79.3 ± 2.4** |
| | TCGA-BRCA[Mol] | 77.9 ± 1.9 | 78.0 ± 1.4 | 78.4 ± 2.5 | 80.8 ± 1.7 | **81.7 ± 2.2** |
| | TCGA-BRCA[OS] | 60.2 ± 4.7 | 60.3 ± 2.9 | 63.3 ± 4.9 | **65.0 ± 6.0** | 64.7 ± 5.7 |
| Colorectal cancer | TCGA-CRC[MSI] | 76.1 ± 4.4 | 88.5 ± 2.5 | 79.7 ± 4.1 | 88.1 ± 1.9 | **91.0 ± 2.2** |
| | TCGA-COAD[OS] | 57.7 ± 10.4 | 59.4 ± 10.2 | 58.3 ± 6.3 | 60.1 ± 10.9 | **62.8 ± 12.7** |
| Lung cancer | TCGA-NSCLC[CType] | 92.8 ± 2.5 | 96.2 ± 1.7 | 94.2 ± 2.8 | 97.3 ± 0.4 | **97.7 ± 1.3** |
| | TCGA-LUAD[OS] | **59.1 ± 4.1** | 55.3 ± 4.8 | 53.7 ± 5.5 | 58.1 ± 4.1 | 53.8 ± 4.5 |
| | TCGA-LUSC[OS] | 59.8 ± 3.7 | 61.6 ± 4.2 | 60.9 ± 5.4 | 60.5 ± 2.1 | **62.2 ± 2.9** |
| Ovarian cancer | TCGA-OV[HRD] | 51.6 ± 4.9 | 69.2 ± 12.9 | 68.0 ± 8.9 | 68.5 ± 10.8 | **74.2 ± 8.6** |
| Kidney cancer | TCGA-RCC[CType] | 97.5 ± 0.8 | 98.6 ± 0.3 | 98.6 ± 0.4 | 98.7 ± 0.3 | **99.5 ± 0.2** |
| Stomach cancer | TCGA-STAD[MSI] | 76.5 ± 3.3 | 78.1 ± 4.8 | 79.6 ± 3.1 | 83.2 ± 8.1 | **89.9 ± 3.9** |
| Pancreatic cancer | TCGA-PAAD[OS] | 59.3 ± 6.8 | 58.2 ± 4.9 | **61.3 ± 2.7** | 57.0 ± 5.5 | 55.3 ± 4.4 |

**Table F2.** Comparison of state-of-the-art SSL frameworks on PAIP-CRC[MSI] external validation after training on TCGA-CRC[MSI] classification task. We display the performance with ABMIL. ROC AUC scores and 95% confidence intervals are computed using bootstrap with 1,000 repeats.

| | | (A) Cohort-specific pre-training | | (B) Pan-cancer pre-training | | |
|---|---|---|---|---|---|---|
| Cancer site | Task | Dino[ViT-S] BRCA | MoCoV2 [RN50W2] COAD | HIPT | CTransPath | iBOT[ViT-B] PanCancer |
| Colorectal cancer | MSI prediction: TCGA-CRC to PAIP | 88.1 [78.1, 99.1] | 94.0 [88.8, 100.0] | 91.6 [85.2, 100.0] | 88.4 [78.2, 100.0] | **94.7** [89.4, 100.0] |

### Appendix G: ROC AUC scores on patch-level classification tasks

This section highlights ROC AUC scores on both patch-level classification tasks: NCT-CRC-HE-7K and Camelyon17-WILDS.

**Table G1.** Comparison of patch classification performance for (A) in-domain pretraining vs out-of-domain training, (B) MoCoV2 vs iBOT frameworks with TCGA-COAD pre-training. ROC AUC scores and $95\%$ confidence intervals are computed using bootstrap with $1{,}000$ repeats. Bold indicates the highest performance across classes.

| | | NCT-CRC-HE-7K | | | | | | | | | Camelyon 17WILDS |
|---|---|---|---|---|---|---|---|---|---|---|---|
| | Method | Adi | Deb | Lym | Muc | Mus | Norm | Str | Tum | All | Metastases |
| (A) | Sup [RN50] IN | 99.8 | 87.1 | **99.9** | 98.5 | **96.3** | 99.8 | 91.6 | 99.5 | 96.5 [96.2, 96.9] | 96.5 [96.3, 96.6] |
| (A) | iBOT [ViT-S] COAD | **100.0** | **99.9** | **99.9** | **100.0** | 93.4 | **100.0** | **97.4** | **99.9** | **98.8** [98.6, 99.0] | **98.3** [98.2, 98.4] |
| (B) | MoCoV2 [RN50W2] COAD | **100.0** | **100.0** | 99.6 | 99.8 | 90.7 | 99.9 | 97.1 | 99.1 | 98.3 [98.0, 98.5] | 97.1 [96.9, 97.3] |
| (B) | iBOT [ViT-B] COAD | 99.9 | **100.0** | **99.8** | **100.0** | **97.4** | **100.0** | **98.7** | **99.9** | **99.5** [99.4, 99.6] | **99.0** [98.9, 99.1] |

**Table G2.** Impact of ViT architecture scaling on patch classification tasks. ROC AUC scores and $95\%$ confidence intervals are computed using bootstrap with $1{,}000$ repeats. Bold indicates the highest performance across classes.

| | NCT-CRC-HE-7K | | | | | | | | | Camelyon 17WILDS |
|---|---|---|---|---|---|---|---|---|---|---|
| Method | Adi | Deb | Lym | Muc | Mus | Norm | Str | Tum | All | Metastases |
| iBOT [ViT-S] COAD | **100.0** | 99.9 | 99.9 | **100.0** | 93.4 | **100.0** | 97.4 | **99.9** | 98.8 [98.6, 99.0] | 98.3 [98.2, 98.4] |
| iBOT [ViT-B] COAD | 99.9 | **100.0** | 99.8 | **100.0** | **97.4** | **100.0** | **98.7** | **99.9** | **99.5** [99.4, 99.6] | **99.0** [98.9, 99.1] |
| iBOT [ViT-L] COAD | 99.9 | 99.9 | **100.0** | **100.0** | **97.4** | **100.0** | 98.3 | **99.9** | 99.4 [99.3, 99.6] | 98.1 [98.0, 98.3] |

**Table G3.** Impact of ViT pre-training datasets on patch classification tasks performance for a ViT-B architecture. ROC AUC scores and $95\%$ confidence intervals are computed using bootstrap with $1{,}000$ repeats. Bold indicates the highest performance across classes.

| | NCT-CRC-HE-7K | | | | | | | | | Camelyon 17WILDS |
|---|---|---|---|---|---|---|---|---|---|---|
| Method | Adi | Deb | Lym | Muc | Mus | Norm | Str | Tum | All | Metastases |
| iBOT [ViT-B] COAD | 99.9 | **100.0** | **99.8** | **100.0** | **97.4** | **100.0** | **98.7** | 99.9 | **99.5** [99.4, 99.6] | 99.0 [98.9, 99.1] |
| iBOT [ViT-B] PanCancer | **100.0** | **100.0** | **99.8** | **100.0** | 95.8 | 100.0 | 98.1 | **100.0** | 99.2 [99.1, 99.3] | **99.5** [99.4, 99.6] |

**Table G4.** Comparison of state-of-the-art SSL frameworks on patch classification tasks. HIPT$^*$[ViT$_{256}$] correspond to the first ViT-S model of HIPT architecture pre-trained on $256 \times 256\,\mathrm{px}$ tiles. ROC AUC scores and $95\%$ confidence intervals are computed using bootstrap with $1{,}000$ repeats. Bold indicates the highest performance across classes.

| | NCT-CRC-HE-7K | | | | | | | | | Camelyon 17WILDS |
|---|---|---|---|---|---|---|---|---|---|---|
| Method | Adi | Deb | Lym | Muc | Mus | Norm | Str | Tum | All | Metastases |
| Dino [ViT-S] BRCA | 99.9 | 99.7 | 99.8 | 99.0 | **98.4** | 99.3 | 89.8 | 99.4 | 98.2 [98.0, 98.3] | 96.6 [96.4, 96.8] |
| MoCoV2 COAD | **100.0** | **100.0** | 99.6 | 99.8 | 90.7 | 99.9 | 97.1 | 99.1 | 98.3 [98.0, 98.5] | 97.1 [96.9, 97.3] |
| HIPT$^*$[ViT$_{256}$] | 99.9 | 99.7 | 99.7 | 99.6 | 97.8 | 99.4 | 92.2 | 99.6 | 98.5 [98.3, 98.7] | 95.4 [95.2, 95.6] |
| CTransPath | **100.0** | 97.1 | **100.0** | 99.9 | 96.9 | 99.9 | 93.9 | 99.7 | 98.4 [98.2, 98.7] | 98.3 [98.1, 98.4] |
| iBOT [ViT-B] PanCancer | **100.0** | **100.0** | 99.8 | **100.0** | 95.8 | **100.0** | **98.1** | **100.0** | **99.2** [99.1, 99.3] | **99.5** [99.4, 99.6] |
